# Single-cell spatial proteomics identifies the JAK/STAT pathway as an actionable therapeutic target in lethal cutaneous drug reactions

**DOI:** 10.1101/2023.11.11.23295492

**Authors:** Thierry M. Nordmann, Holly Anderton, Akito Hasegawa, Lisa Schweizer, Peng Zhang, Pia-Charlotte Stadler, Ankit Sinha, Andreas Metousis, Florian A. Rosenberger, Maximillian Zwiebel, Takashi K. Satoh, Florian Anzengruber, Maria C. Tanzer, Yuki Saito, Ting Gong, Marvin Thielert, Haruna Kimura, Natasha Silke, Edwin H. Rodriguez, Restivo Gaetana, Hong Ha Nguyen, Annette Gross, Mitchell P. Levesque, Peter J. Murray, Saskia Ingen-Housz-Oro, Andreas Mund, Riichiro Abe, John Silke, Chao Ji, Lars E. French, Matthias Mann

**Affiliations:** Department of Proteomics and Signal Transduction; Max Planck Institute of Biochemistry; Martinsried, Bavaria, 82152; Germany; Department of Dermatology, University Hospital Zurich, University of Zurich, Zurich; Switzerland; Inflammation division, Walter and Eliza Hall Institute of Medical Research, Melbourne, VIC, Australia; Department of Medical Biology, University of Melbourne, Parkville, VIC 3052, Australia; Division of Dermatology, Niigata University Graduate School of Medical and Dental Sciences, Niigata, Japan; Department of Dermatology, The First Affiliated Hospital of Fujian Medical University, Fuzhou, Fujian, China; Department of Dermatology and Allergy, University Hospital, Ludwig Maximilian University (LMU) Munich, Germany; Department of Internal Medicine, Division of Dermatology, Cantonal Hospital Graubuenden, Chur, Switzerland; Advanced Technology and Biology division, Walter and Eliza Hall Institute of Medical Research, Melbourne, VIC, Australia; Immunoregulation Research Group, Max Planck Institute of Biochemistry, Martinsried, Germany; Dermatology Department, AP-HP, Henri Mondor Hospital, Créteil, France; Proteomics Program, The Novo Nordisk Foundation Center for Protein Research, University of Copenhagen, Faculty of Health and Medical Sciences, Blegdamsvej 3B, 2200 Copenhagen, Denmark; Key Laboratory of Skin Cancer of Fujian Higher Education Institutions, Fujian Medical University, Fuzhou, Fujian, China; Dr. Philip Frost, Department of Dermatology and Cutaneous Surgery, University of Miami Miller School of Medicine, Miami, FL

## Abstract

Toxic epidermal necrolysis (TEN) is a fatal drug-induced skin reaction and an emerging public health issue. Triggered by common medications, TEN patients undergo severe and sudden epidermal detachment caused by keratinocyte cell death. Although molecular mechanisms driving keratinocyte cell death have been proposed, the main drivers remain unknown and no effective therapy exists. To systematically map molecular changes that are associated with TEN and identify potential druggable targets, we employed the single-cell spatial proteomics technique Deep Visual Proteomics. We analyzed formalin-fixed paraffin-embedded archived skin-tissue biopsies of three types of cutaneous drug reactions with varying severity and quantified over 5,000 proteins in keratinocytes and skin-infiltrating immune cells. Most strikingly, this revealed a robust enrichment of Type-I and -II interferon signature in the immune cell and keratinocyte compartment of TEN patients, along with a drastic activation of pSTAT1. Targeted inhibition with pan-JAK inhibitor (JAKi) tofacitinib reduced keratinocyte-directed cytotoxicity in a novel live-cell imaging assay, using patient-derived keratinocytes and peripheral blood mononuclear cells (PBMCs). Furthermore, oral administration of pan-JAKi tofacitinib or baricitinib ameliorated clinical and histological disease severity in two distinct mouse models of TEN. Lastly, JAKi treatment was safe and associated with rapid cutaneous re-epithelialization and recovery in four patients with TEN. This study uncovers the JAK-STAT and interferon signaling pathways as key pathogenic drivers of TEN and demonstrates the potential of targeted JAK inhibition as a curative therapy.

## Introduction

Cutaneous adverse drug reactions (CADRs) are the most common immune-mediated adverse drug reactions, with up to 2% of CADRs classified as severe and life-threatening^1^. The spectrum of CADRs ranges from self-resolving maculopapular rash (MPR) to rare and life-threatening conditions, such as drug reaction with eosinophilia and systemic symptoms (DRESS) and toxic epidermal necrolysis (TEN)^2^. DRESS is a severe multiorgan hypersensitivity reaction triggered by culprit drugs and sustained by the reactivation of latent viruses, leading to aberrant T-cell and eosinophilic response^3^. In comparison, TEN is characterized by fulminant epidermal detachment of over 30 % of the body surface area and is fatal in a third of all cases. TEN represents a considerable socio-economic burden due to specialized in-hospital care and lost productivity among survivors^4–6^. Despite diverse proposed pathogenic mechanisms^7–21^, the main drivers of cytotoxicity remain elusive. Accordingly, consensus therapy of TEN is still supportive care^22^.

Over the last 5 years, spatial omics has emerged as a powerful strategy to obtain a global, molecular view of intact tissue samples^23–25^. When applied at single-cell resolution, these techniques provide an opportunity to profile different cells without removing them from their native environment and thus provide a more physiologically relevant view of cellular function and dysfunction. However, although a broad variety of such technologies exist, translation into direct clinical benefit has remained largely aspirational. For example, application of spatial omics to TEN has been limited by the rarity of severe cutaneous drug reactions, and inability of existing techniques to explore different skin-residing cellular subsets especially in the context of clinically relevant formalin-fixed paraffin-embedded (FFPE) skin biopsies. To address these technical limitations, we have recently introduced Deep Visual Proteomics (DVP) as a novel spatial single-cell proteomics technology that combines the use of high content imaging, AI guided cell segmentation and classification, and laser micro dissection of individual target cells to achieve ultra-sensitive proteomics^26^. By using FFPE-archived biopsies, DVP enables high-yield, in-depth analysis of cells of interest, using easily accessible tissue specimens, even in rare disorders. Here, we employed DVP to study mechanisms of cutaneous drug reactions, investigate the predominant molecular features in the severe types of CADRs, and functionally validate targeted small molecule inhibitors for rapid therapeutic intervention in the most severe CADR, TEN.

### Cohort assembly and spatial proteomics workflow

To study molecular mechanisms of cutaneous drug reactions, we assembled an age and sex matched, retrospective cohort of lesional FFPE skin biopsies from patients with mild (MPR) or severe (TEN, DRESS) CADRs alongside healthy controls (Fig. 1a and Extended Data Figure 1, N = 21). After clinical and histological re-validation of all cases, we stained 3 μm FFPE tissue sections for CD45 to visualize immune cells and with pan-cytokeratin for keratinocytes (Fig. 1b). A machine learning (ML) algorithm ensured proper segmentation and prevented downstream analysis of overlapping cellular compartments (Extended Data Fig. 1b). Thereafter, contours of immune cells and keratinocytes from each patient were laser micro- dissected and each separately pooled into multi-well plates for subsequent peptide extraction and ultra-high sensitivity mass spectrometry (MS)-data acquisition (Fig. 1b). This DVP approach achieved a proteomic depth of 4992 unique proteins in immune cells and 5009 in keratinocytes across all patients (Fig. 1c). The biological separation of keratinocytes from immune cells greatly exceeded interpatient heterogeneity (Fig. 1c and Extended Data Fig. 1c). A third of all identified proteins were differentially expressed, including the upregulation of LGALS7B and KRT5 in keratinocytes and of ANXA6 and CORO1A in immune cells, confirming the cell-type resolution of the DVP data (Extended Data Fig. 1d). Taken together, our cohort, which included 5-6 individuals per CADR indication, was amenable to the DVP workflow, resulting in robust and reproducible quantification of ∼5,000 proteins and clear dissection of keratinocytes and immune cells.

**Fig. 1.**
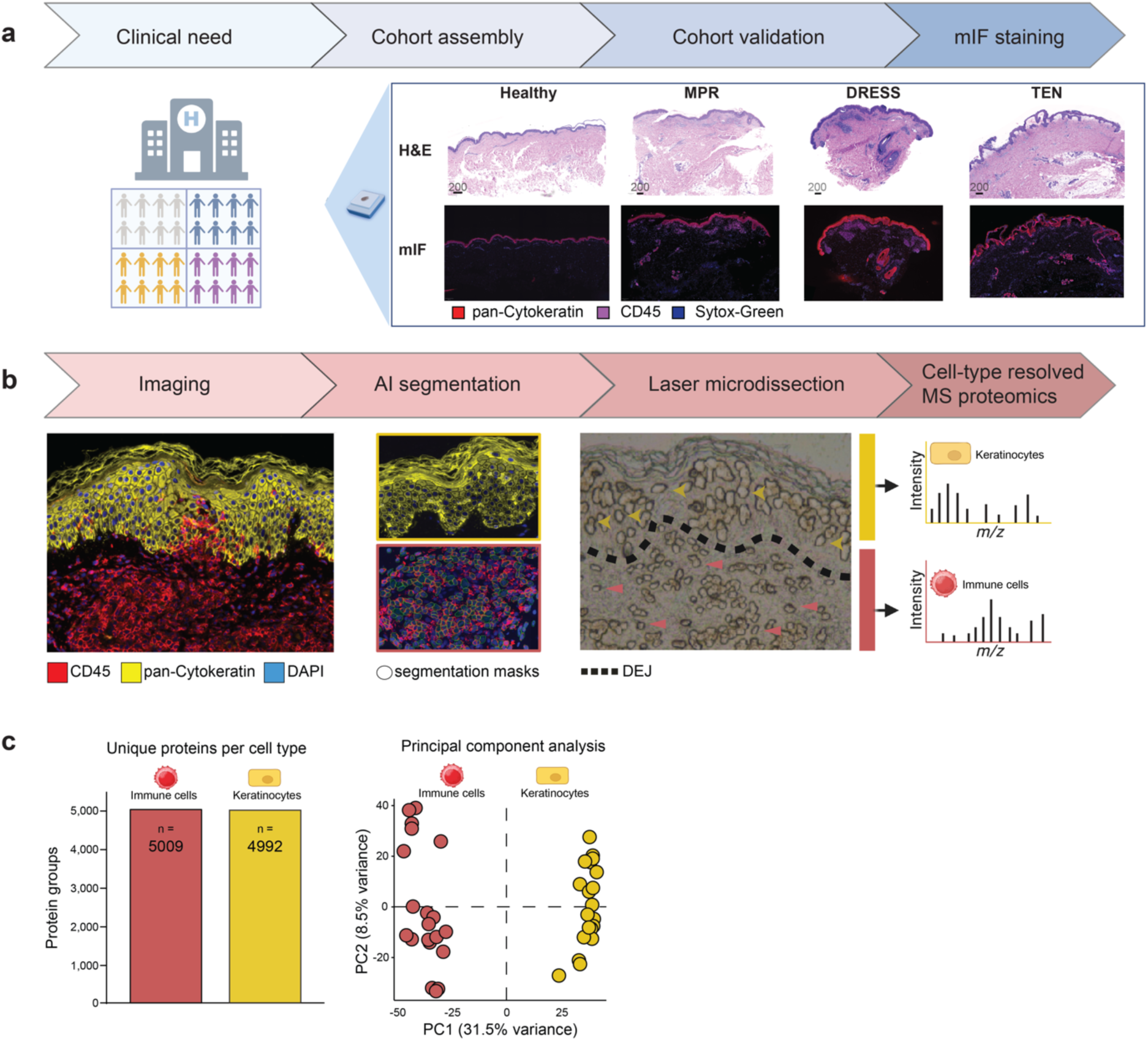
Deep Visual Proteomic workflow for in-depth proteomic data with cellular resolution in cutaneous adverse drug reactions. **a.** Cohort assembly and validation (N=21). Hematoxylin & Eosin (H&E) and multicolor immunofluorescence staining of a representative 3μm FFPE tissue section from the indicated conditions (red: pan-cytokeratin^+^ keratinocytes, purple: CD45^+^ immune cells, blue: sytox-green stained nuclei). Maculopapular rash (MPR), drug rash with eosinophilia and systemic symptoms (DRESS), toxic epidermal necrolysis (TEN) and healthy individuals. N = 5 individuals per cohort (TEN, DRESS, MPR and healthy). **b.** The Deep Visual Proteomics pipeline, including high-content imaging, machine learning-based cell segmentation and classification, followed by laser microdissection and ultra-sensitive mass spectrometry-based proteomics at cellular resolution. Representative area of excised keratinocyte (yellow arrow) and immune cell (red arrow) from FFPE tissue sections. **c.** Identification of unique protein groups and principal component analysis across all patients in the indicated cell type. Each dot is a patient (N = 21), color represents the cell type. This figure was partially created with Biorender.com.

### The proteome of lesional keratinocytes in cutaneous drug reactions

Having acquired the cell-type resolved proteomes of our cohort, we performed comprehensive data analysis of lesional keratinocytes of the selected CADRs. We quantified a median of 3788 protein groups across all patients, covering a dynamic range of 4.2 orders of magnitude (Fig. 2a and Extended Data Fig. 2a). Data completeness and quantitative accuracy were excellent (median coefficient of variation (CV) of 17.4% per cohort, despite patient heterogeneity, Extended Data Fig. 2b-c). Keratinocyte proteomes clustered according to the type of drug reaction, with Principal Component 1 separating TEN from other CADRs and keratinocytes from healthy individuals (Fig. 2b). PC1 was enriched in proteins involved in calcium homeostasis and cellular stress response, including HSPA5 (heat-shock protein family A member 5), TMCO1 (transmembrane and coiled coil domains 1) and ATP2A2 (ATPase sarcoplasmic/endoplasmic reticulum Ca^2+^ transporting 2; Fig. 2c). Interestingly, STAT3 (signal transducer and activator of transcription 3) was also amongst the main drivers of PC1, congruent with a dominant interferon signature in gene set enrichment analysis of PC1 (GSEA; Fig. 2c, d). Pairwise comparison of diseased to healthy proteomes revealed that the number of differentially expressed proteins (DEP) increased with the severity of the CADRs (Extended Data Fig. 2d). Thus, MPR, the mildest CADR, had no DEP when compared to healthy, whereas both DRESS and TEN exhibited significant changes. DRESS robustly upregulated multiple MHC class-I proteins in keratinocytes, suggesting that keratinocytes participate actively in culprit-drug associated antigen presentation (TAP1, TAP2, TAPBP, HLA-A; log_2_ fold change > 1, Fig. 2e).

**Fig. 2.**
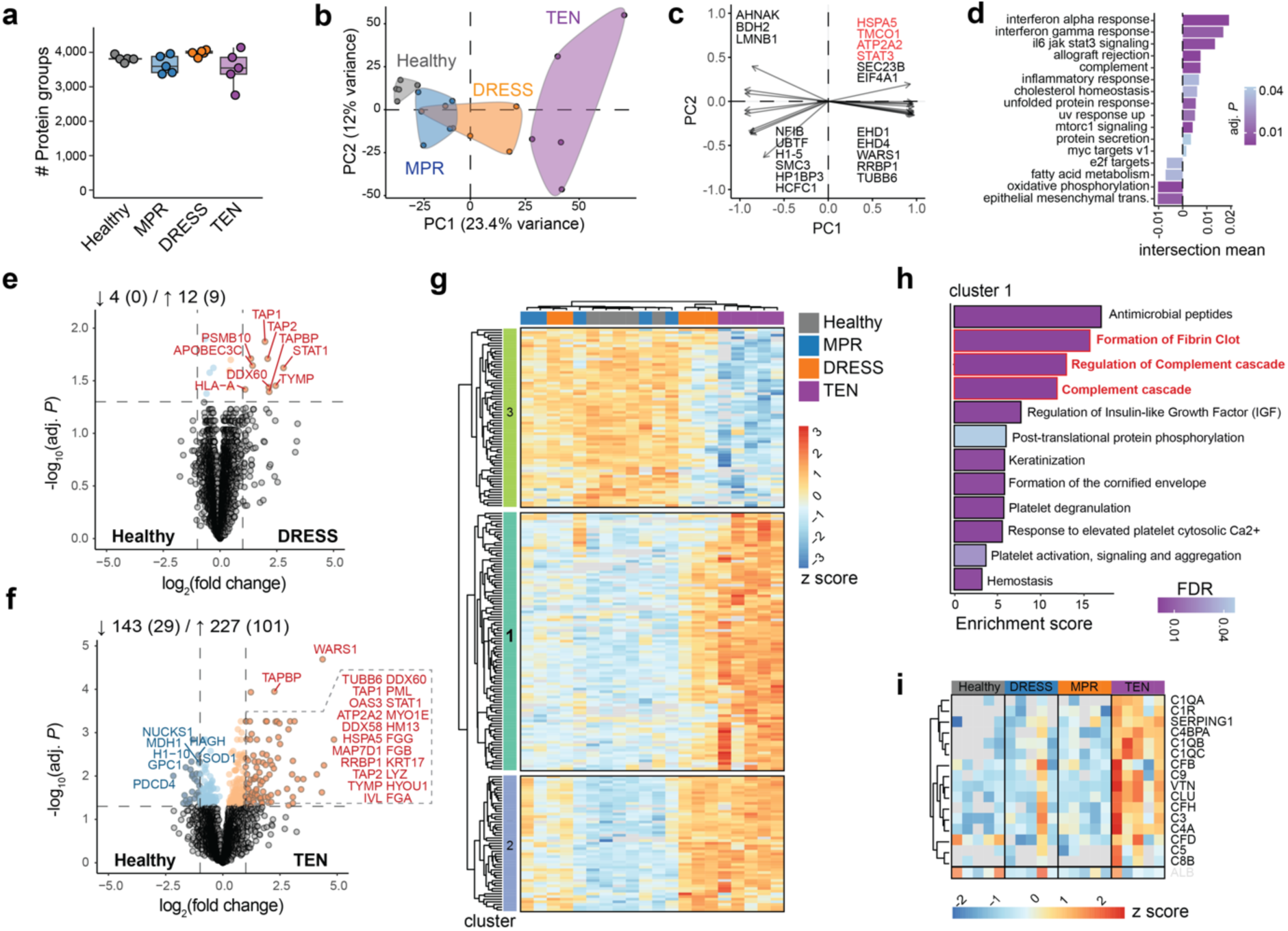
**a**. Number of proteins identified in keratinocytes across cohorts. Box plots show the median (center line) with interquartile range of 25% to 75% and 95% confidence interval. **b.** Principal Component (PCA) analysis, main drivers of PC1/2 separation (**c**) and PC1 gene set enrichment analysis (**d**). **e-f.** Differentially expressed proteins (DEPs) between the indicated condition and healthy. Colored dots are significant (BH adjusted *P* < 0.05, FDR 0.05), dashed vertical line represents log_2_ fold change of ≤ 1 or ≥ 1. **g.** Unsupervised hierarchical clustering with analysis of variance (ANOVA) significant proteins (adjusted *P* < 0.05, post-hoc Tukey HSD). Color indicates normalized intensities (z score) **h.** Overrepresentation analysis of ANOVA cluster 1, ordered according to enrichment score. Color indicates degree of significance. **i.** Semisupervised heatmap of complement factors in the indicated conditions, and albumin (at the bottom) to exclude serum contamination, color indicates normalized values (z score). N = 5 individuals per cohort, mean of biological duplicates per patient.

In keratinocytes from TEN patients, the most severe CADR, there were 227 up - and 143 significantly down-regulated proteins, including those involved in antimicrobial response, such as lysozyme and clusterin (Fig. 2f). WARS1, one of the most strongly upregulated proteins (log_2_ fold change 4.36, *adj. P* 2.04 x 10^-5^), has not previously been associated with TEN. This tryptophanyl-tRNA synthetase has a non-canonical role in activating the innate immune system as an endogenous TLR2/4 ligand, thereby triggering cytokine and chemokine production^27^. Expression of superoxide-dismutase (SOD) 1 was reduced by half. This is interesting because oxidative stress and reactive oxygen species (ROS) upregulate keratinocyte Fas-ligand expression, a key cytolytic molecule involved in keratinocyte apoptosis and epidermal detachment in TEN^17,28^. Unsupervised hierarchical clustering of ANOVA significant proteins between all three CADR types, showed a clear alignment for TEN and three distinct protein clusters, revealing a further degree of stratification (Fig. 2g). Overrepresentation analysis of cluster 1, which represents proteins with highest intensity values in TEN, revealed multiple terms involved in the regulation of complement activation and blood clotting (Fig. 2h and Extended Data. Fig. 2e-f). In accordance, nearly all complement factors in our keratinocyte proteome dataset are uniquely upregulated in keratinocytes of TEN (Fig. 2i and Extended Data. Fig. 2g). Taken together, proteomes of keratinocytes derived from severe CADRs, DRESS and TEN, reveal distinct disease-associated protein expression signatures.

### The proteome of lesional immune cells in cutaneous drug reactions

Cutaneous immune cell infiltration in TEN is thought to be sparse and scattered, in contrast to the dense and confined perivascular infiltrates in MPR and DRESS^29^. To understand underlying molecular differences in immune cells of the skin in CADRs, we assessed the in-depth proteome of immune cells (median of 3418 per patient, median CV 19.8% within cohorts, with high data completeness; Fig. 3a and Extended Data Fig. 3a-c). These proteomes again clustered according to disease phenotype and were separated from healthy controls, but in contrast to keratinocytes, Principal Component 1 of immune cells separated DRESS from other CADRs (Fig. 3b and Extended Data Fig. 3d). Gene set enrichment analysis of PC1 was prominently associated with targets of E2F and MYC (Extended Data Fig. 3e). Proteins driving this include RNA processing and splicing factors such as DDX39A (DExD-Box Helicase 39A), EIF4A3 (Eukaryotic Translation Initiation Factor 4A3) and SRSF6 (Serine and Arginine Rich Splicing Factor 6), indicating a highly proliferative state (Extended Data Fig. 3 f). PC1 and PC3 together revealed unique features of DRESS and TEN. In DRESS, multiple viral pathways terms are enriched, in line with the postulated role of latent viral reactivation in its pathogenesis (Fig. 3c). In contrast, a dominant interferon signature was evident in TEN, together with STAT1 as a main driver (Fig. 3c and Extended Data Fig. 3f). In direct proteomic comparison to healthy control, DRESS had the highest number of DEPs (n = 730), followed by TEN (n = 233) and MPR (n = 128; Fig. 3d - g). DEPs unique to TEN included GCA (Grancalcin, log_2_ fold change 1.56, q 2.38 x 10^-2^) and PDIA3 (Protein Disulfide Isomerase Family A Member 3, log_2_ fold change 0.642, q 2.51 x 10^-2^) (Fig. 3f), both not previously described in the context of TEN. Similar to its role in cancer^30^, PDIA3 may modulate MHC-class I and subsequent CD8^+^-driven cytotoxicity in TEN.

**Fig. 3.**
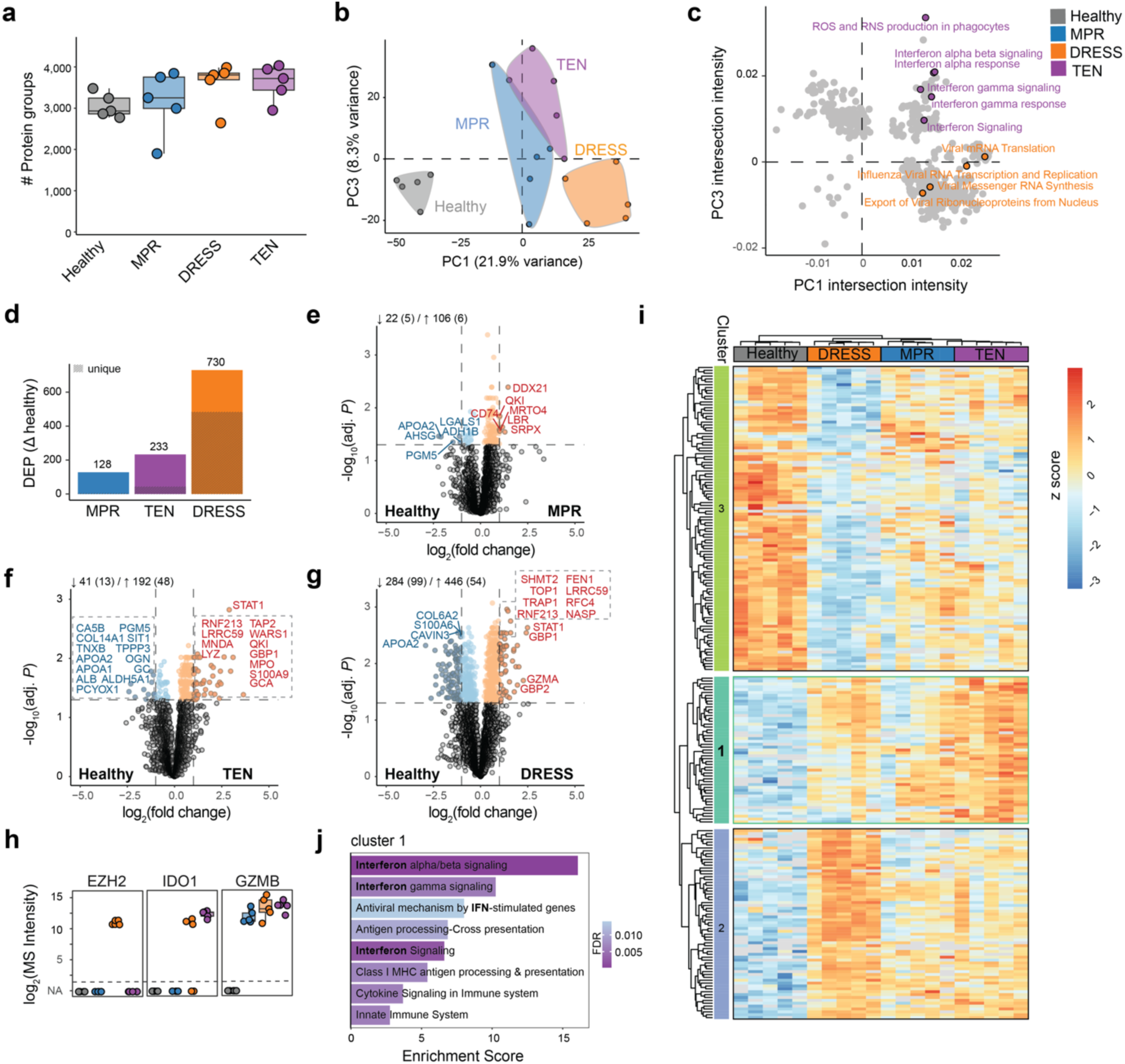
**a.** Number of proteins identified in immune cells across all cohorts. Box plots show the median (center line) with interquartile range of 25% to 75% and 95% confidence interval. **b.** Principal Component (PCA) analysis of the proteomic results. **c.** Intersection of PC1 and PC3 gene set enrichment analysis. Terms including “viral” or “interferon” are highlighted. **d-g.** DEPs in the indicated conditions versus healthy and corresponding volcano plots. Shaded area within columns represents the proportion of unique proteins. In the volcano plots, color represents significance (*adj. P* < 0.05), dashed vertical line represents log_2_ fold change of ≤ 1 or ≥ 1. **h.** Selected proteins of interest, with a distinct identification across the cohorts **i.** Unsupervised hierarchical clustering of ANOVA significant proteins (*adj. P* < 0.05, post-hoc Tukey HSD). Color indicates normalized intensities (z score). **j.** Overrepresentation analysis of ANOVA cluster 1, ordered according to enrichment score. Color indicates degree of significance (FDR). N = 5 individuals per cohort, mean of biological duplicates per patient.

A minority of proteins were specifically identified within defined CADR subsets (Extended Data Fig. 3g). Of particular interest is histone methylase EZH2 (Enhancer of Zeste homolog), the catalytic member of *PRC* (Polycomb Repressive Complex) 2, detectable only in immune cells from DRESS patients (Fig. 3h). Immunofluorescence staining, as well as near-exclusive transcription in a data set of PBMC-derived lymphocyte cluster from a therapy resistant DRESS patient^31^, confirmed our proteomic observation (Extended Data Fig. 3h - i). In fact, EZH2 fold change over healthy control was higher than the main targets reported in that study (STAT1, JAK3, IL2RG)^31^. While the role of EZH2 in cancer prompted the development of an FDA-approved inhibitor, its contribution to inflammatory and allergic diseases has not been extensively investigated^32^.

IDO (Indoleamine 2, 3-dioxygenase 1) was only detected in severe CADRs and represents a further protein of interest with an already available, clinically approved inhibitor (Fig. 3h). Whether IDO1 has a detrimental role or dampens the cytotoxic reaction, as within the tumor microenvironment^33^, requires further investigation. Although granzyme B has been described as a key mediator of cytotoxicity in TEN^21^, our proteomics data showed that it is present at similar levels in non-cytotoxic DRESS (Fig. 3h).

Unsupervised hierarchical clustering of ANOVA significant proteins in immune cells resulted in three visually distinct protein clusters with perfect disease alignment (Fig. 3i). Overrepresentation analysis of Cluster 2, which mainly represents DRESS, confirmed the highly replicative features of infiltrating immune cells in this disease, illustrated by multiple terms for DNA replication (Extended Data Fig. 3j). Its most prominent term (Activation of ATR in response to replication stress) was dominated by the DNA replication factor MCM (Minichromosome Maintenance protein complex) and RFC (Replication Factor C; Extended Data Fig. 3k). Intriguingly, human MCM associates with EBV replication origin (oriP) for the propagation of the viral genome^34^. This potentially serves as a direct link to the viral pathogenesis of this disorder. Importantly, proteins in cluster 1, which dominantly represents TEN and are only mildly elevated in MPR and DRESS, were almost exclusively associated with type I and II interferon signaling (Fig. 3j). Given the urgent clinical need for a specific therapy for TEN, we further focused on the role of the interferon pathway in TEN.

### The JAK/STAT pathway is drastically activated in TEN

Severe inflammatory disorders are often characterized by feedback loops that exacerbate the immune response^35^. We therefore hypothesized that proteins significantly regulated in both keratinocytes and immune cells would identify therapeutically relevant pathways. We identified six such proteins (WARS1, STAT1, S100A9, LYZ, GBP1 and APOL2) that were all at least four-fold upregulated in both keratinocytes and immune cells of TEN compared to healthy control (Fig. 4a). Strikingly, all six proteins are part of the interferon pathway, responsible for signal transduction (STAT1) or triggered by it^36–40^. Interferons signal through the Janus Kinase / Signal Transducer and Activator of Transcription (JAK/STAT) and we examined the molecular components of this signaling cascade using our comprehensive cell-type resolved proteomic dataset (Fig. 4b and Extended Data Fig. 4a-b). STAT1 and STAT3 were upregulated in both immune cells and keratinocytes, while STAT2 was up in keratinocytes and STAT5 in immune cells. JAKs have low abundance and the only one we identified was JAK3 in TEN. Most downstream interferon-stimulated genes (ISG) were either strongly upregulated, illustrated by a 9-fold increase of GBP1, or only detectable in TEN (e.g. GBP1). This points to a strong upregulation of all components of the JAK-STAT pathway in TEN.

**Fig. 4.**
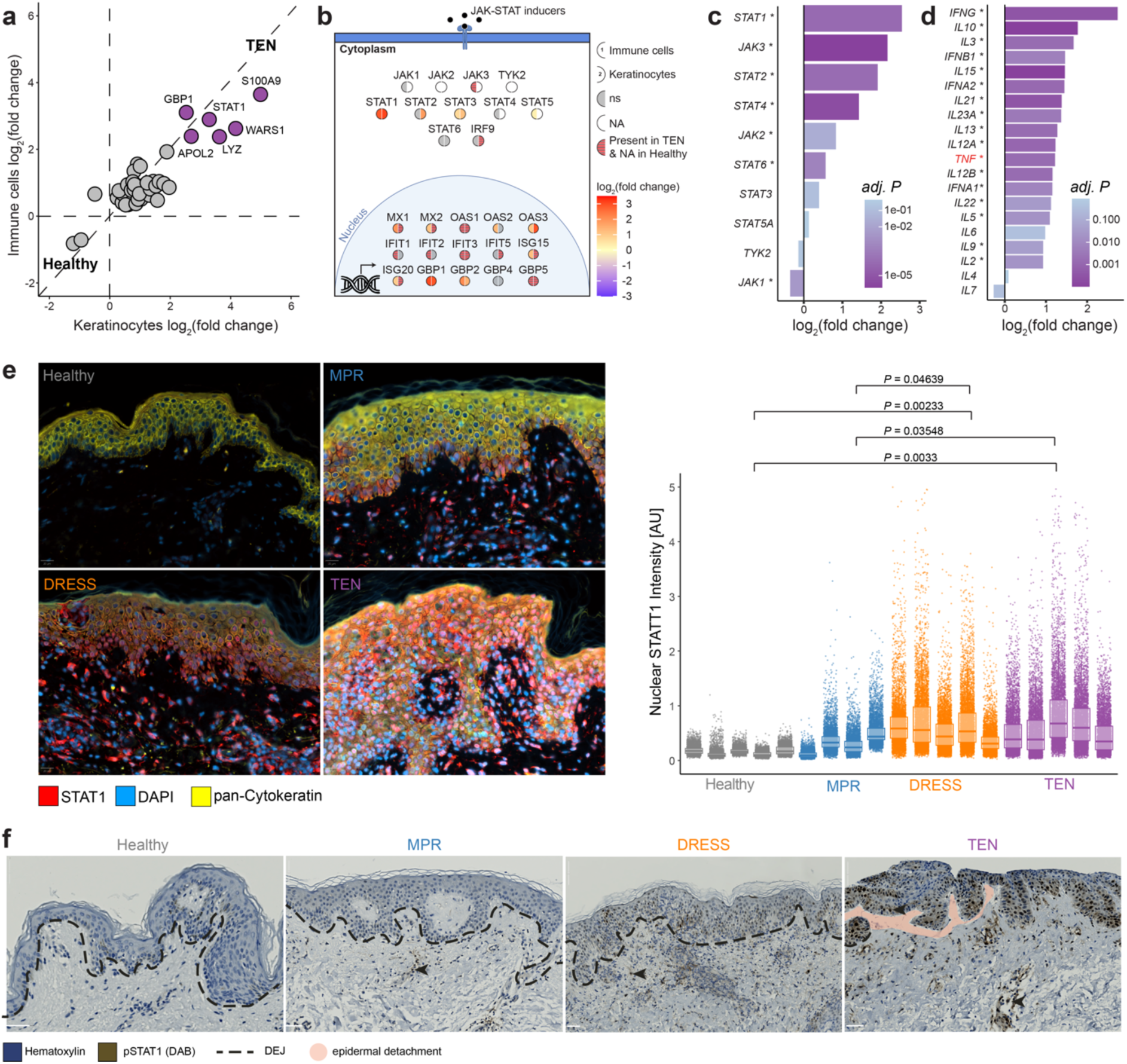
**a.** Significantly quantified keratinocyte (x-axis) and immune cell (y-axis) proteins and their corresponding log_2_ fold change compared to healthy. Purple color indicates proteins with log_2_ fold change > 2 in both cell types. Dashed vertical and horizontal lines indicate fold change = 0 and the dashed diagonal line represents equal regulation. **b.** Schematic illustration of quantified JAK/STAT pathway proteins in both cell types. Color represents log_2_ fold change compared to healthy, with direct comparison between immune cells (left semi-circle) and keratinocytes (right) of the indicated protein. Empty (semi-)circles are proteins with missing values. All colors except for grey are significantly regulated (*adj. P* < 0.05). **c.** mRNA expression levels in TEN compared to healthy for the indicated genes of the JAK/STAT pathway (validation cohort) **d.** mRNA expression change in TEN compared to healthy of genes signaling through the JAK/STAT pathway (validation cohort). Asterisks indicate significance (*adj. P* < 0.05). **e.** Representative immunofluorescent staining of FFPE tissue sections against Stat1 (red) and pan-Cytokeratin (yellow). Quantification of nuclear Stat1 intensity per cell, normalized to its own nuclear staining intensity, for all samples of the corresponding conditions. Box plots show the median (center line) with interquartile range of 25% to 75%. **f.** Representative phospho-STAT1 immunohistochemistry staining. Dashed lines represent the dermo-epidermal junction (DEJ). Epidermal detachment is highlighted with red (in TEN). N = 5 individuals per cohort (TEN, DRESS, MPR and healthy) and mean of biological duplicates per patient, except for **c** and **d** where N = 10 per cohort (TEN, healthy).

We confirmed substantial and broad JAK/STAT activation in TEN and also in Steven-Johnson- Syndrome (SJS)-TEN overlap by transcriptomic analysis in a second CADR cohort (N=20, Fig. 4c and Extended Data Fig. 3k). SJS-TEN overlap is defined by an affected body surface area of 10 – 30% in contrast to over 30% in TEN, with an otherwise identical phenotype. Of cytokines known to signal through the JAK/STAT pathway, interferon-gamma gene expression was the most strongly upregulated in TEN, exceeding that of cytokines such as TNF, known to be involved in TEN^41^ (Fig. 4d). Visualization of STAT1 in FFPE tissue sections further confirmed its significant upregulation in TEN (Fig. 4e). Notably, the degree of single-cell nuclear STAT1 upregulation in TEN was similar to DRESS, in which therapeutic intervention with JAK-inhibitor proved to be successful^31^. To further establish STAT1 activation in TEN at the signaling level, we proceeded with histological analysis of phosphorylated STAT1. This revealed a profound degree of STAT1 phosphorylation in both skin-residing immune cells and keratinocytes of TEN patients (Fig. 4f). Collectively, these results suggest that activation of the JAK/STAT pathway is a key driver of TEN.

### The JAK/STAT pathway is an actionable therapeutic target in preclinical models of TEN

To investigate the clinical significance of our experimental findings, we developed a novel *in-vitro* model that authentically replicates key aspects of cutaneous drug reactions (Fig. 5a). To this end, we cultivated primary keratinocytes from single hair follicles of TEN survivors, building on previous protocols^42^. Concurrently, we isolated patient-matched PBMCs and activated them using CD3/28. After five days, we fluorescently labeled the keratinocytes and co-cultured them with activated or resting PBMCs. We introduced a fluorescent DNA-binding dye to identify cells with compromised membrane integrity, in order to visualize, track and quantify the cytotoxic action of activated PBMCs towards keratinocytes using live-cell imaging. In this autologous co-culture model, keratinocytes were efficiently killed within 72 hours in the presence of activated PBMCs (Fig. 5b). Applying the pan-JAK inhibitor tofacitinib, an FDA and EMA approved drug, resulted in dose-dependent inhibition of keratinocyte cytotoxicity, whereas co-culturing of keratinocytes with resting PBMCs or exposure of keratinocytes to tofacitinib alone did not induce keratinocyte cell death (Fig. 5c).

**Fig. 5.**
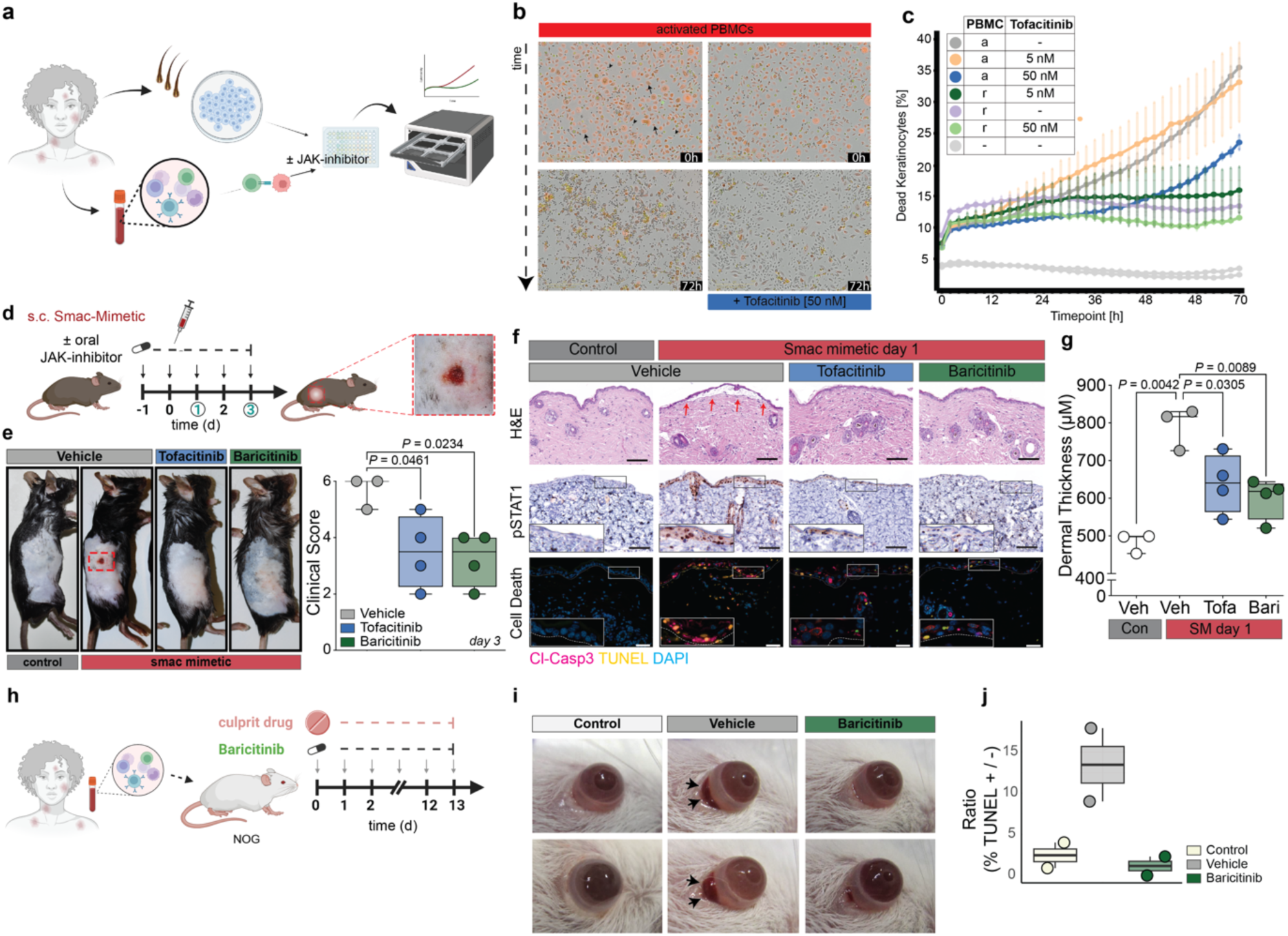
**a.** Live-cell imaging co-culture assay with keratinocytes and activated PBMCs from the same patient ± JAK inhibitor, to assess keratinocyte cell death **b.** Keratinocytes labeled with CellTracker Red (arrow) in co-culture with unlabeled PBMCs (arrowhead) at timepoint 0 and 72h, ± 50nM tofacitinib. **c.** Kinetic data of **b.** quantified over time (a = activated; r = resting; n=6 replicates per condition). **d.** Experimental protocol to assess efficacy of oral JAK inhibitor tofacitinib (30 mg/kg) or baricitinib (10 mg/kg) in combination with s.c. SMAC mimetics, which cause a TEN phenotype (red arrows: epidermal detachment). **e.** Representative macroscopic image and clinical assessment score to evaluate the macroscopic effect of treatment of the corresponding cohort on day three (n = 3 for vehicle and 4 for tofacitinib or baricitinib group). **f.** Representative H&E, pSTAT1 and cell death (magenta = cleaved caspase 3 / yellow = TUNEL / cyan = DAPI) microscopy image of the corresponding cohort. Inserts magnify indicated area of interest. Scale bars = 100μM. **g.** Average dermal thickness of the indicated conditions on day 1 (each data point is a mouse, N=15 measurements/mouse). **h.** Experimental protocol to assess efficacy of baricitinib (10 mg/kg) in a humanized mouse model of TEN, using patient PBMCs. **i.** Ocular reaction (black arrows) following daily culprit drug administration ± oral baricitinib (10 mg/kg). **j.** Ratio of TUNEL positive over TUNEL negative subepithelium of the corresponding cohort (n = 2 per condition) Box plots show the median (center line) with interquartile range of 25% to 75% and 95% confidence interval.

Encouraged by these results, we next evaluated the efficacy of oral JAK inhibitors in established mouse models of TEN^43,44^. Loss of Inhibitor of Apoptosis proteins (IAPs) shifts TNF signaling towards apoptosis^45^ and necroptosis^46^. Both cell death modalities contribute to the pathology of TEN^19,47^. In mice, subcutaneous (s.c.) injection of a small molecule IAP antagonist drug (smac-mimetic, SM) induced TEN-like cutaneous inflammation, epidermal detachment and histological features of TEN (Fig. 5d-f). Oral treatment with JAKi (tofacitinib 30 mg/kg or baricitinib 10 mg/kg) markedly reduced macroscopic disease severity on day 1 and 3 as quantified by a clinical assessment score composed of all major TEN hallmarks (Nikolsky sign, edema, rubor; Fig. 5e and Extended Data Fig. 5a). Similar to human TEN, a strong increase in pSTAT1 was detectable following SM injection and reduced disease severity through treatment with JAK inhibitors was accompanied by a significant reduction in cutaneous pSTAT1 levels (Fig. 5f, and Extended Data Fig. 5b). Average dermal thickness was elevated upon SM injection compared to vehicle and restored to normal levels upon JAKi (Fig 5g and Extended Data Fig. 5c). Further, JAKi also decreased keratinocyte cell death (TUNEL^+^/cleaved caspase-3 (CC3)^+^, Fig. 5f and Extended Data Fig. 5d,) and showed less immune cell infiltration (CD45^+^, Extended Data Fig. 5b). Interestingly, in both inhibitor groups CC3 positive cells were more frequently TUNEL negative indicating an earlier stage of the apoptotic program (Fig. 5f). This suggests, that the process of cell death is dampened and delayed as a result of JAK inhibition. Importantly, epidermal recovery following s.c. SM injection was accelerated in JAKi treated mice, demonstrated by the strong expression of the proliferation marker Ki67 in the basal layer, ruling out a detrimental effect of treatment on wound healing (Extended Data Fig. 5b).

Despite the striking similarities of the SM-induced murine model with human TEN, SM primarily engages downstream pathways which represent a potential limitation in translating these findings to the human context. To address this, we further tested the efficacy of JAK inhibitor baricitinib in an established humanized mouse model of TEN, in which patient PBMCs of a TEN survivor are injected intravenously into immunocompromised NOD/Shi-scid, IL-2Rγnull (NOG) mice (Fig. 5h). Following daily oral administration of the causative drug (acetaminophen) severe ocular conjunctivitis and cell death of conjunctival epithelium reminiscent of TEN occurred at day 14 (Fig. 5i – j and Extended Data Fig. 6a - b and d-e)^19^. In this second mouse model of TEN, baricitinib treatment reduced (clinical) ocular conjunctivitis and (histological) cell death of conjunctival epithelium as compared to vehicle treatment (TUNEL^+^/TUNEL^-^ ratio for baricitinib: 1.15% (mean) ± 1.63 (SD) and Vehicle: 13.45% (mean) ± 6.29 (SD); Fig. 5j and Extended Data Fig. 6a - g). Taken together, the *in vitro* and *in vivo* data clearly demonstrates the efficacy of JAKi in preclinical models of TEN.

### Patients with toxic epidermal necrolysis recover after JAK inhibition

Based on our preclinical data and the urgent clinical need in this devastating disease, we treated four patients with TEN or SJS/TEN overlap with off-label JAKi treatment. One such patient was receiving carboplatin, etoposide and the PD1-antagonist serplulimab for small-cell lung cancer (Fig. 6). After the third cycle of anti-cancer treatment, he developed TEN with epidermal detachment affecting 35% of his body surface and a severity-of-illness score (SCORTEN^48^) of 4, indicating a predicted mortality rate of 58.3%. Epidermal detachment progressed despite high-dose intravenous methylprednisolone (80 mg d^-1^) and therefore we initiated JAK1 inhibitor rescue therapy with abrocitinib, an acknowledged drug in the field of dermatology and recently approved for treatment of atopic dermatitis^49^. Within two days, the disease stopped progressing and re-epithelialization was visible within four days, reaching near-completeness after 16 days (95%), after which the patient was discharged following full recovery from TEN. All four patients treated with JAKi survived at day 30 and no side effects occurred (Extended Data Fig. 7a - d). Notably, cutaneous pSTAT1 levels were drastically reduced in all patients following JAK-inhibitor treatment (Extended Data Fig. 7e). This clinical data shows the potential of repurposing approved JAK1 inhibitor abrocitinib or pan-JAK inhibitors such as tofacitinib for the treatment of TEN.

**Fig. 6.**
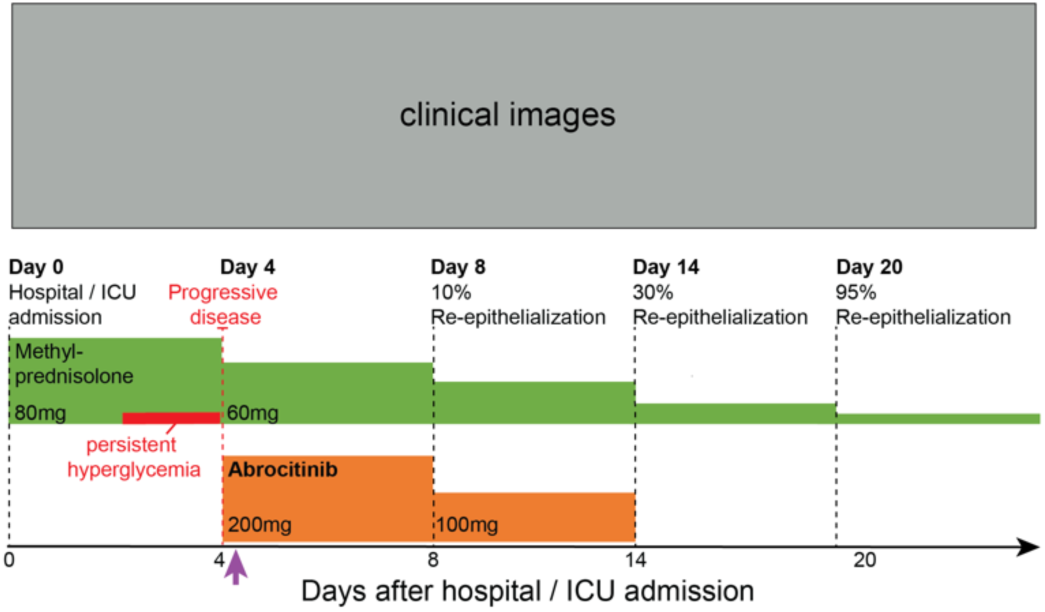
Disease course under JAK1 inhibition (abrocitinib) in a severe case of toxic epidermal necrolysis. Arrow marks timepoint starting with abrocitinib.

## Discussion

The skin is the most frequently affected organ by adverse drug reactions, and these reactions occur in 1 – 3 % of all hospitalized patients^50^. Severe CADRs and, in particular, TEN have an elusive pathogenesis and cause substantial fatality^2^. Here, we used deep spatial proteomic profiling of keratinocytes and immune cells to analyze FFPE tissue sections from patients exhibiting three different types of CADR and healthy donors, and quantified ∼5,000 proteins in both types of cells. This in-depth proteomic analysis across all major types of cutaneous drug reactions with cell-type resolution provided unique insights into the molecular mechanism of each CADR and identified targets for therapeutic interventions of these disorders. Skin tissue is composed of many different cell types and Deep Visual Proteomics allowed a nuanced view of these cell-type specific proteomic changes, even in rare samples with sparse and scattered immune cell infiltration. For instance, the striking upregulation of WARS1 in keratinocytes and immune cells of TEN is of special interest, in light of the increasingly appreciated role of the innate immune system in this disease^51^. Both immune cells and keratinocyte carry functional TLR receptors, including TLR2 and 4^52^. Thus, our results may link the non-canonical role of WARS1 as an endogenous TLR2/4 ligand to the immune pathogenesis of TEN. Given its ability to trigger cytokine and chemokine production, it would be interesting to investigate if secreted WARS1 directly contributes to the damage occurring in TEN by initiating and exacerbating the immune response.

In skin-infiltrating immune cells of DRESS, the most dominant finding was the appearance of the histone methyltransferase EZH2, which we further confirmed by histology. EZH2 plays a key role in epigenetic gene regulation by modifying histone H3 at lysine 27 (H3K27me3). Functional consequences mainly attribute to the increased proliferation of cancer cells; however, recent studies suggest a more nuanced, quantitative effect on defining cellular states^53^. EZH2 is the third most common site of mutation (29%) in patients with idiopathic hypereosinophilic syndrome^54^. Intriguingly, systemic eosinophilia is a hallmark of DRESS and the primary culprit of organ damage, indicating a potential connection to EZH2 dysfunction^55^. In addition, E2F pathway enrichment detected in our proteomic dataset correlates with the overexpression of EZH2, which functions as a downstream effector in this signaling cascade^56^. Together, these findings strongly suggest that EZH2 and, by extension, epigenetic modifications, may play a significant part in the pathogenesis of DRESS. As EZH2 is inhibited by the FDA-approved drug tazemetostat, elucidating the precise role of EZH2 in DRESS could open new therapeutic avenues for this severe, multiorgan inflammatory CADRs^57^.

The major finding of our study, is the drastic upregulation of the JAK/STAT pathway in TEN, which is not observed in less severe CADRs. Our results pinpoint this signaling pathway as the main driver of cutaneous inflammation, keratinocyte cytotoxicity and epidermal detachment. Our cell-type resolved findings indicate a more active role of keratinocytes than previously anticipated in driving the skin directed cytotoxic immune response. Many components of the JAK/STAT signaling cascade are upregulated in both immune cells and keratinocytes of TEN, highlighting the potential for detrimental self-amplifying nature of interferon signaling that could explain the extensive tissue damage^58^.

To date, standard of care for TEN is limited to immediate cessation of the causative drug and best supportive care, usually in an intensive care unit. Unfortunately, under these standard of care conditions hospital mortality rates range from 15-20 percent, highlighting the urgent need for more effective treatments. Our spatial proteomics results nominated the JAK/STAT pathway as an actionable therapeutic target in TEN, and we provided *in vitro* and *in vivo* preclinical evidence that JAK inhibitors substantially reduced keratinocyte-directed cytotoxicity.

Importantly, we treated four patients with TEN or SJS-TEN overlap syndrome, two of which were previously unresponsive to high-dose systemic corticosteroids. All four showed a remarkable response to JAK inhibitors and were discharged in good health after treatment. Taken together, our data strongly suggest that JAK inhibition is feasible and beneficial in TEN. Given the high short-term risk of mortality associated with TEN and the short duration of specific therapy required, it appears that the potential benefits of short-term use of JAK inhibitor therapy, if confirmed in larger clinical trials, could outweigh the potential risks of long-term JAK inhibition observed in patients with chronic inflammatory disorders^59^. Due to strong association of developing TEN during immune checkpoint inhibition (OR= 4.33)^60^, JAKi may be especially attractive to rapidly resume oncological treatment in case this adverse reaction develops. While our study with JAK-inhibition in a small number of patients cannot be considered conclusive in terms of efficacy and should, therefore, be interpreted with caution, its potential for the management of TEN, a life-threatening disease with no efficient treatment so far, should be taken into consideration.

These data pave the way for a clinical trial of early JAKi therapy in TEN with the potential to change the outcome of this lethal adverse reaction to common drugs. Our study is also a successful example of directly linking emerging cell-type resolved spatial omics technologies, especially spatial proteomics, with a new treatment modality that is of benefit to patients. We envision that similar approaches could be transformative in a broad set of inflammatory or oncological conditions by helping to target treatments more precisely.

## Data Availability

All data produced in the present study will be available after peer-review, when published.

## Acknowledgements

We thank all members of the Proteomics and Signal Transduction group at the Max Planck Institute of Biochemistry, especially Dr. Maximilian T. Strauss for bioinformatic cell segmentation strategies, Katharina Zettl for superb immunofluorescence staining and Bianca Splettstoesser for cell culture. We thank Dr. Katrin Kerl-French for revalidation of all cases. Dr. Leonie Zeitler and Dr. Patricia P. Ogger provided technical assistance for the live-cell imaging experiments. The J2 / 3T3 cells were a generous gift from Prof. Hans-Dietmar Beer (University Hospital Zurich, Switzerland).

## Author contributions

T.M.N., L.E.F. and M.M. conceived the project. T.M.N., H.A. A.H., R.A., J.S., L.E.F. and M.M. designed the experimental framework. T.M.N. performed the proteomic experiments with help from L.S., A.M., M.Z. and M.T. H.A., M.C.T., N.S. and J.S., performed and analyzed the smac mimetic mouse model experiments. A.H., Y.S., H.K., H.H.N. and R.A. performed and analyzed the humanized mouse model experiments. P.Z., T.G. and C.J. performed and analyzed the JAK inhibition in patients. T.M.N., H.A., J.S., L.E.F. and M.M. designed the figures with help from F.A.R., M.T. and L.S.. T.M.N. performed the live-cell imaging experiments and analyses, with substantial experimental guidance from A.G. and P.J.M. Transcriptomics experiments and analyses were performed by P.C.S. and T.K.S., whereas L.S., A.S., A.M., F.A.R., M.Z., M.T., E.H.R. and A.M. provided intellectual input and helped interpret data. F.A., R.G., M.P.L. and S. I-H-O. provided intellectual input and helped construct the cohorts. T.M.N., L.E.F. and M.M. wrote the manuscript. All authors read, revised and approved the manuscript.

## Methods

### Patient biopsies

Skin tissue biopsies were obtained during routine histopathological diagnostic procedures at the Departments of Dermatology and Allergology of the University Hospital Zurich (proteomic cohort) and Ludwig Maximilian University of Munich (validation cohort). In the proteomic cohort, TEN patients had a minimal affected skin area of 30%. In the validation cohort, patients with SJS/TEN overlap were included (minimal affected skin >10%). All patients with DRESS had a RegiSCORE of 6-7 (definite diagnosis). In addition to the typical clinical features, the lymphocyte transformation test (LTT) or skin-tests were positive in all MPR patients. Histopathological diagnosis was re-validated in all cases by a board-certified dermatopathologist using a fresh H&E-stained tissue section. Baseline characteristics were statistically evaluated using Analysis of Variance (ANOVA) for numeric variables (age) and a Chi-squared test of independence for categorical variables (sex). All obtained disease biopsies were performed in the context of routine clinical workup for retrospective analysis with informed consent and ethical approval in place (Munich: 22-0342, 22-0343; Zurich: BASEC: Req-2021-00226 and 2017--00494; Fujian: MRCTA, ECFAH of FMU[2023]400. All experiments were performed in accordance with the Declaration of Helsinki.

### Treatment of patients with JAK inhibitors

Regular assessment and vigilant surveillance of the vital signs, hematological parameters and coagulation markers was performed throughout treatment. Patients with active infections were excluded. All patients were additionally treated according to current best supportive care guidelines^22^. Oral abrocitinib was administered at a regimen of 200mg daily for 5-7 days, and tapered to 100mg daily for 5-7 days. Similarly, oral tofacitinib was administered at a dosage of 10mg daily for 5-7 days, and then tapered to 5mg daily for 5-7 days. In cancer patients with high SCORTEN, a reduction in JAKi dosage was be initiated after observing a certain degree of re-epithelialization. Treatment was approved by the local ethics committee and the institutional review board of the First Affiliated Hospital of Fujian Medical University (Fujian: MRCTA, ECFAH of FMU[2023]400), and the patient was provided written informed consent.

### Mice

#### Smac mimetic TEN mouse model

As previously published^43^ male mice were injected subcutaneously with 100μl of 1mg/ml smac mimetic (SM) CompA (TetraLogic Pharmaceuticals) or vehicle (12% Captisol). JAK inhibitor (10 mg/kg Baricitinib or 30 mg/kg Tofacitinib; Selleckchem) or vehicle (20% Captisol; 50μl; Selleckchem) was administered by oral gavage twice daily starting one day before s.c. SM injection. Mice were sacrificed on either day one or day three post SM injection, the site was photographed, scored, and samples collected for ex-vivo analysis. We used a four-point ordinal scale (0-4) to assess the three clinical associated macroscopic criteria of TEN (Oedema, rubor and epidermal disruption (Nikolksy sign / lesion severity)) at day one or three post SM injection. They were assessed and scored as 0 (not. present), 1 (mild), 2 (moderate), or 3 (severe) and the scores were combined for an overall clinical score. Experiments were approved by the local ethics committee (WEHI AEC# 2022.009).

#### Humanized mouse model of TEN

Immunocompromised NOD/Shi-scid, IL-2Rγnull (NOG) mice at 6 weeks of age were included in the experiment with n = 2 in each of Baricitinib treatment and both control groups. PBMCs (2×10^6^) from a patient who had recovered from SJS/TEN 1 year earlier were injected intravenously into the NOG mice, followed by oral administration of the causative drugs (acetaminophen, 1.5 mg/100 μl) at day one and once daily thereafter. The dosage used in this model was based on mg / kg body weight, converted from the normal adult human dose. Baricitinib (10 mg/kg) or vehicle (20% Captisol; 50 μl) was administered by oral gavage twice daily from day one. Body weight, ocular reactions or cutaneous changes were assessed daily. On day 14, the degree of conjunctivitis was evaluated after ocular dislocation under general anesthesia and scored (0 = no conjunctivitis, 1 = mild conjunctivitis, 2 = severe conjunctivitis). The ratio of the number of TUNEL-positive cells to the total conjunctival cell count in a 400x field was calculated for all mice. The patients received no systemic glucocorticoids at the time of PBMCs collection. Experiments were approved by the local ethics committee and the institutional review board of Niigata University, and the patient was provided written informed consent.

### Membrane slide tissue mounting, immunofluorescence staining, imaging and laser microdissection

2 μm PEN membrane slides (MicroDissect GmbH) were exposed to UV light (254 nm) for one hour and then coated with Vectabond (Vector laboratories; SP-1800-7) according to the manufacturers protocol. 3 μM thick tissue sections of each FFPE block were mounted on the pretreated membrane slides and dried overnight at 37°C. Immunofluorescence was performed according to our previously optimized protocol for membrane slides^61^. In brief, slides were heated to 56°C for 20min and immediately deparaffinized / rehydrated (Xylol 2x2 min, 100% EtOH 2x1 min, 90% EtOH 2x1 min, 75% EtOH 2x1 min, 30% EtOH 2x1 min, ddH_2_0 2x1 min). Slides were then transferred to prewarmed glycerol-supplemented Antigen Retrieval buffer^61^ (DAKO pH9 #S2367 + 10% Glycerol) at 88°C for 20 min, followed by a 20 min cooldown at RT. Slides were then washed in water and blocked with 5% BSA / PBS for 30 min, followed by overnight primary antibody incubation at 4°C in a humid staining chamber (1:100 mouse monoclonal CD45, DAKO M0701; 1:200 rabbit polyclonal CK Pan, DAKO Z0622). After washing in PBS, secondary antibodies (1:400 anti-mouse IgG A647, Invitrogen A32728; 1:400 anti-rabbit IgG A555, Invitrogen A32732) were incubated for 1 h at RT. After washing in PBS, sections were counterstained with SYTOX™ Green (1:400, Invitrogen S7020) for 10 min, washed again and coverslipped using Slowfade Diamond Antifade Mountant (Invitrogen, S36963). Sections were imaged on a Zeiss Axioscan at 20x magnification with a tile overlap of 10%. At an excitation wavelength of 553 nm and 631 nm, 100% laser intensity were used, while 50% was used in the 493 nm channel. Illumination time was adapted to the optimal spectral properties. Five z-stacks at an interval of 1.25 μm were acquired. Multi-scene images were then split into single scenes, z-stacks combined to a single plane by extended depth of focus (variance method, standard settings) and stitched, using the proprietary Zeiss Zen Imaging software. Images were imported as .czi files into the Biological Image Analysis Software (BIAS) with the packaged import tool^26^. Singe-cell segmentation was performed using deep neural network on the basis of pan-cytokeratin for keratinocytes and CD45 for immune cells at 1.0 input spatial scaling, 50% detection confidence and 30% contour confidence. Only contours between 30 μm^2^ and 200 μm^2^ were taken into consideration. After removal of duplicates at tile-overlapping regions, a supervised machine learning approach was used to remove false identifications and overlapping cell types. Contours were exported together with three calibration points that were chosen at characteristic tissue positions. Contour outlines were simplified by removing 99% of data points. For keratinocytes, only every second shape was (randomly) chosen in order to prevent membrane instability while cutting multiple adjacent cells. Contour outlines were imported after reference point alignment, and shapes were cut by laser microdissection with the LMD7 (Leica) with a 63x objective in a semi-automated manner at the following settings: power 57, aperture 1-2, speed 23, middle pulse count 1, final pulse -3, head current 46 - 53%, pulse frequency 2600, offset 210. We collected 700 contours for each keratinocyte sample and 1000 for each immune cell sample. For each patient, biological duplicates were collected when possible, from the same slide at different regions. Dissected contours were collected directly into the same underlying low-binding 384-well plate (Eppendorf 0030129547), immune cells in the top half of the plate and keratinocytes in the bottom half, leaving an empty well between each patient. The plate was sealed with adhesive foil (Covaris), centrifuged at 1,000xg for 1 minute and then kept at RT until further processing.

### Sample preparation for downstream LC/MS

Semi-automated sample preparation and digestion was performed in the collection plate using the Bravo pipetting robot (Agilent) as previously described^26^. For this, wells were washed with 28 μl of 100% acetonitrile and dried in a SpeedVac for 20 min at 45°C. The contours were then resuspended in 4 μl of 60mM triethylammonium bicarbonate (TEAB, Sigma) in MS-grade H2O, sealed with two adhesive foils and heated at 95°C for 60min in a 384-well thermal cycler (Eppendorf). 1 μl of 60% acetonitrile in 60 mm TEAB (12.5% final acetonitrile concentration) was added, followed by heating at 75°C for 60 min in a 384-well thermal cycler. Samples were then predigested with 4 ng of Trypsin for 4 h followed by overnight digestion with 6 ng LysC in a 384-well thermal cycler at 37°C. After 18 h the reaction was quenched with 1.5 µl 6% trifluoroacetic acid (1% final concentration). Samples were then manually transferred to single PCR tubes, dried in a SpeedVac for approx. 60 min at 45°C and stored at -20°C.

### LC-MS/MS analysis

Peptides were resuspended in 4.1μl MS loading buffer (2% AcN (v/v) + 0.1% TFA (v/v) in MS-grade H20) immediately prior to measurement. All samples were stratified into cell-type and replicates, and further randomized using the “rand” function in Excel. An EASY nanoLC 1200 (Thermo Fisher Scientific) was coupled to a timsTOF SCP mass spectrometer (Bruker) via a nanoelectrospray ion source (Captive spray source, Bruker). Peptides were separated on a 50 cm in-house packed HPLC column (75 um inner diameter, 1.9 um ReproSil-Pur C18-AQ silica beads (Dr. Maisch GmbH)), which was heated to 60°C by an in-house manufactured oven. A linear gradient of 120 min was ramped at a constant flow rate of 300 nl/min from 3 to 30% buffer B in 95 min, followed by an increase to 60% for 5 min, washed at 95% buffer B for 10 min and re-equilibration for 10 min at 5% buffer B (buffer A: 0.1% formic acid (FA) and 99.9% ddH2O; buffer B: 0.1% FA, 80% ACN, and 19.9% ddH2O). The timsTOF SCP mass spectrometer was operated in dia-PASEF mode using the standard 16 dia-PASEF scan acquisition scheme^62^ with 4 IM steps per dia-PASEF scan, covering a m/z range from 400 to 1200 and ion mobility of 0.6 to 1.6 Vs cm^-2^. All other settings were standard as described in^63^

### Raw data analysis with DIA-NN

A library-free search was performed, using a DL predicted spectral library in DIA-NN (v1.8.0)^64^. Uniprot human databases UP000005640_9606 and UP000005640_9606_additional were used. MS raw files from immune cells and keratinocytes were searched separately in DIA-NN, apart from data shown in Figure 1c. Methionine oxidation was defined as variable modification and missed cleavages were limited to one. The precursor charge ranged from 2 to 4, precursor mass range was set to 300 to 1,800, and peptide length from 7 to 35. Mass and MS1 accuracy were set to 15, based on prior estimation. Isotopologues, and MBR were enabled and neural network classifier was set to single-pass mod. The ‘--relaxed-prot-inf’ function was activated using the command line for more conservative protein grouping. Proteins were inferred from FASTA. Library generation was set as ‘Smart profiling’, ‘RT-dependent’ as cross-run normalization an ‘Robust LC (high precision)’ as quantification strategy.

### Preparation and subsequent transcriptomic analysis of FFPE tissue sections

Ten-micrometer formalin-fixed, paraffin embedded human skin sections were cut from each sample and deparaffinized using ROTICLEAR® (Carl Roth GmbH + Ko. KG). RNA was isolated using RecoverAllTM Total NucleicAcid Isolation Kit (AM1975, Thermo Fisher Scientific) and concentrated using the Savant™ SpeedVac™ DNA 130 Integrated Vacuum Concentator (Thermo Fisher Scientific). Concentration was measured using Nanodrop One/Onec (Thermo Fisher Scientific). 150 µg of RNA from each sample was hybridized over night with unique probe pairs for 624 genes, including 594 genes from the nCounter® Immunology Panel, 15 internal reference genes, and 30 user defined genes using the Panel Plus option (NanoString Tecnhologies). Data were collected using the nCounter® SPRINTTM Profiler (NanoString Tecnhologies).

### Bioinformatics data analysis on MS and transcriptomic data

Bioinformatics data analysis was carried out using the R statistical computing environment version 4.0.2. For subsequent statistical analysis, protein or gene intensities were log2- transformed and average intensities of the biological replicates were computed for each patient and cell type. Aside from the unique protein numbers mentioned in Fig. 1c, only proteins that exhibited an identification rate of 70% or higher in at least one cohort were included, for each cell type independently. Box plots show the median (centre line) with interquartile range of 25% to 75% and 95% confidence interval. For Principal component analysis (PCA), missing values were imputed patient-wise based on a normal distribution (width of 0.3, downshift of 1.8). For the analysis of variance (ANOVA), the R stats package was used; variables were zero-centered and scaled prior to analysis. Subsequent multiple testing correction (*adj. P*) was conducted using the Benjamini-Hochberg (BH) method with a false discovery rate (FDR) cutoff of 5%; Post-hoc Tukey Honest Significant Differences (Tukey-HSD) were calculated at a confidence level of 0.95. For differential expression, the non-paired t-test was performed using the rstatix package with standard settings, followed by multiple comparison correction (BH, FDR 5%) from the stats package. For the comparison between TEN and healthy for selected proteins in both cell types (shown in Figure 4b), a non-paired t-test was performed and the log_2_ fold change value of TEN over healthy was visualized in the figure using a consistent range and color-coding for both cell types. The fill color of the (semi-circles) was manually adjusted using the “eyedropper” tool in Adobe illustrator. Proteomic overrepresentation analyses (ORA) and gene set enrichment analysis (GSEA) were performed with WebGestalt 2019^65^ in the same R environment. The heatmaps were generated using the pheatmap package, with the data being zero-centered and scaled before display.

### Generation of hair follicle-derived keratinocytes for co-culture assay

Hair follicles were placed on thin-Matrigel (Corning, #354234) coated T25 flasks. 1 ml of conditioned mouse embryonic fibroblast medium, from irradiated J2-3T3 cells, was carefully added. Medium was changed every 48 hours. When outgrowth of keratinocytes became visible, medium was aspirated and replaced with 2 ml Keratinocyte medium. When confluent, keratinocytes were detached using TrypLE™ Express Enzyme.

### Isolation of patient PBMCs for co-culture assay

PBMCs were isolated using Ficol according to standard protocol. Cells were frozen in liquid nitrogen until further usage. Keratinocyte expansion and PBMC isolation were approved by the local ethics committee with written informed consent (23-0544).

### Live-cell imaging co-culture assay

Patient PBMCs (2 x 10^5^ / well / 200 μl) were stimulated in U-shape 96-well plates in the presence or absence of CD3/28 (Gibco Dynabeads, #11161D) at a ratio of 1:1, in X-VIVO 15 (Lonza, #881026) for 5 days at 37 °C / 5% CO^2^. On day 4 of PBMC stimulation, hair-follicle derived keratinocytes from the same patient were plated on 96-well plates. On day 5 of PBMC stimulation, keratinocytes were fluorescently labeled (Celltracker Red; Invitrogen #C34552) according to the manufacturer’s recommendation. Activated or non-activated (“resting”) PBMCs were then added to the keratinocytes at a ratio of 1:1, after magnetic removal of CD3/28 beads where necessary. Tofacitinib (CST, #14703) or control (DMSO) was added in the indicated concentration as well as Celltox Green (Promega, G8731, 1:8000 final dilution) to reach a final volume of 200 μl per well. Every 2 hours images were acquired using a live cell analysis system (Incucyte Sx3, Sartorius) for 72 hours at 37 °C / 5% CO^2^. Quantification of dead keratinocytes (Celltox^+^Celltracker^+^ / total Celltracker^+^) was performed using the proprietary analysis software (Incucyte, Sartorius).

### Immunostaining on regular glass slides

3 μm FFPE tissue sections were mounted on regular superfrost-plus glass slides and exposed to high temperatures for improved tissue adherence. Subsequently, sections were deparaffinized and subjected to heat-induced antigen retrieval and blocking solution. The primary antibody was applied to the tissue section for one hour at room temperature or overnight at 4°C (pSTAT1 and cleaved caspase-3). For immunofluorescence staining, a species-specific secondary antibody coupled to a fluorophore was added for an additional hour, followed by a nuclear counterstain. Alternatively, immunohistochemical staining was conducted using horseradish peroxidase (HRP) coupled secondary antibodies, or signal amplified using biotinylated secondary antibodies followed by labelling with and Avidin-Biotin Conjugate (ABC) HRP Kit. HRP labelled samples were then color development with 3,3’-Diaminobenzidine (DAB) and Hematoxylin counterstain. TUNEL staining was performed according to the manufacturer’s instruction prior to application of the anti-cleaved caspase-3 primary antibody for co-staining. H&E staining were performed according to standard protocol. Images were acquired on a Zeiss Axioscan 7 or a DP72 microscope and cellSens imaging software (Olympus), or entire slides were scanned with a 3D Histech Virtual Slide Microscope (Olympus), viewed using CaseCenter (3D Histech, Hungary) and image captured on a MacBook Pro 13-inch retina display (Apple, CA, USA) (Ki67, CD45, H&E).

### Quantification of STAT1 expression in IF images

3 μm FFPE tissue sections of all patients from the proteomic cohort were subjected to multiplex immunofluorescence staining for STAT1, pan-Cytokeratin and Hoechst. All stainings were then uniformly acquired as .czi files using the Zeiss Axioscan 7. Single-cell segmentation was performed within QuPath (v0.4.1) using the standard nuclear segmentation algorithm. After segmentation, all available features were extracted, including staining intensities for all channels and nuclei. Segmentation and feature extraction was automated with QuPath using the script editor, to ensure equality. In R (v. 4.0.2) 5000 nuclei and their corresponding measurements were then randomly chosen per patient and merged into a single file, excluding samples with fewer than 5000 cells (n = 1). To account for staining variation between slides, mean STAT1 fluorescence intensity was divided by its own mean Hoechst intensity on a per-cell basis. Statistical significance between cohorts was determined by a two-tailed t-test on mean normalized STAT1 values. Data is visualized using ggplot2.

