## Extended Data for "Single-cell spatial proteomics identifies the JAK/STAT pathway as an actionable therapeutic target in lethal cutaneous drug reactions"

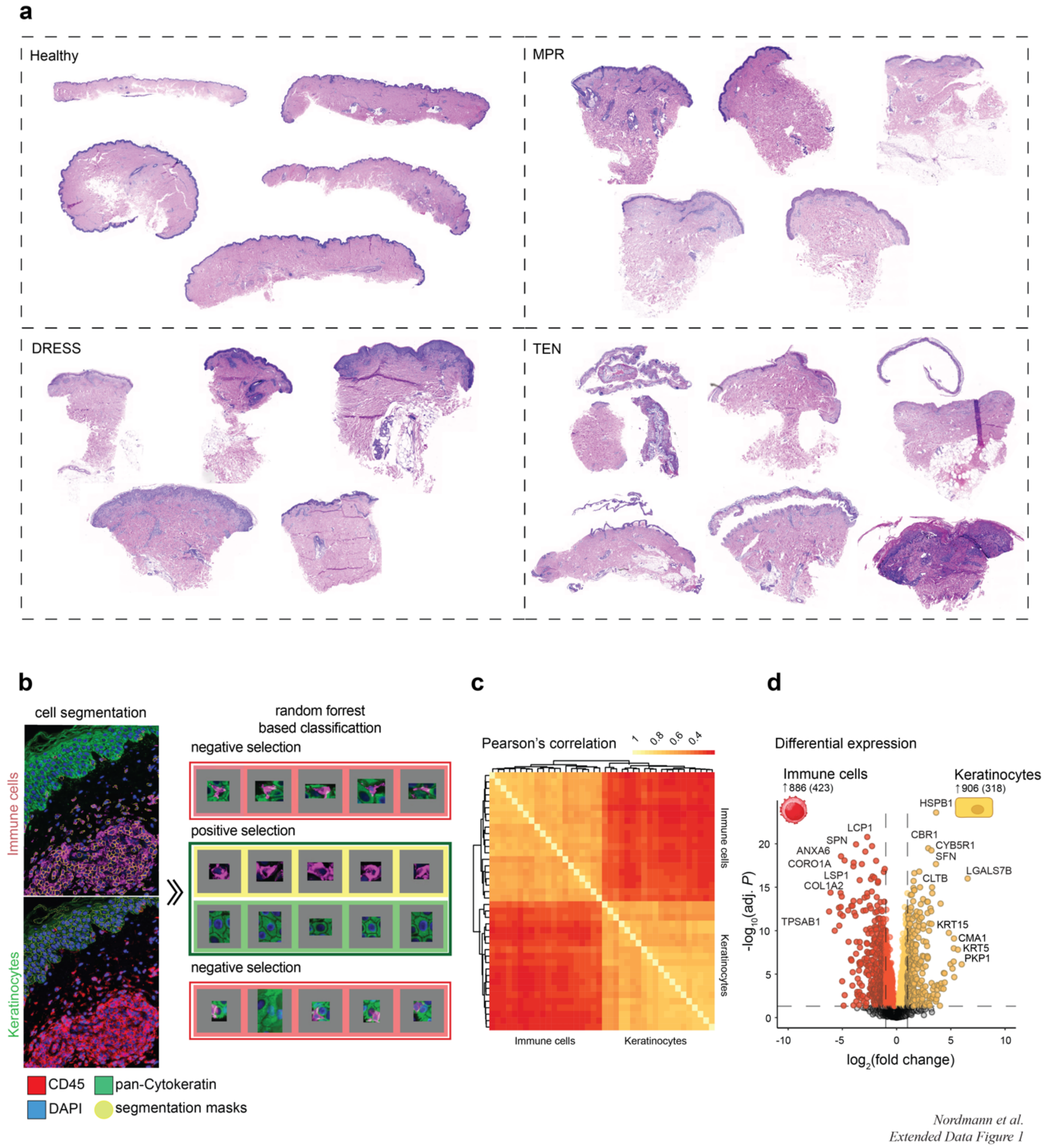


**Extended Data Fig. 1 | a.** Hematoxylin & Eosin staining of all tissue sections of the indicated conditions used for the proteomic cohort. **b.** Machine learning based image segmentation and cell-type classification. **c.** Pearson correlation and **d.** Differentially expressed proteins (DEP) quantified across all patients (N = 21) in the indicated cell type. Colored dots are significant (BH adjusted $P < 0.05$, FDR 0.05), dashed line represents log2 fold change of $\leq 1$ or $\geq 1$.

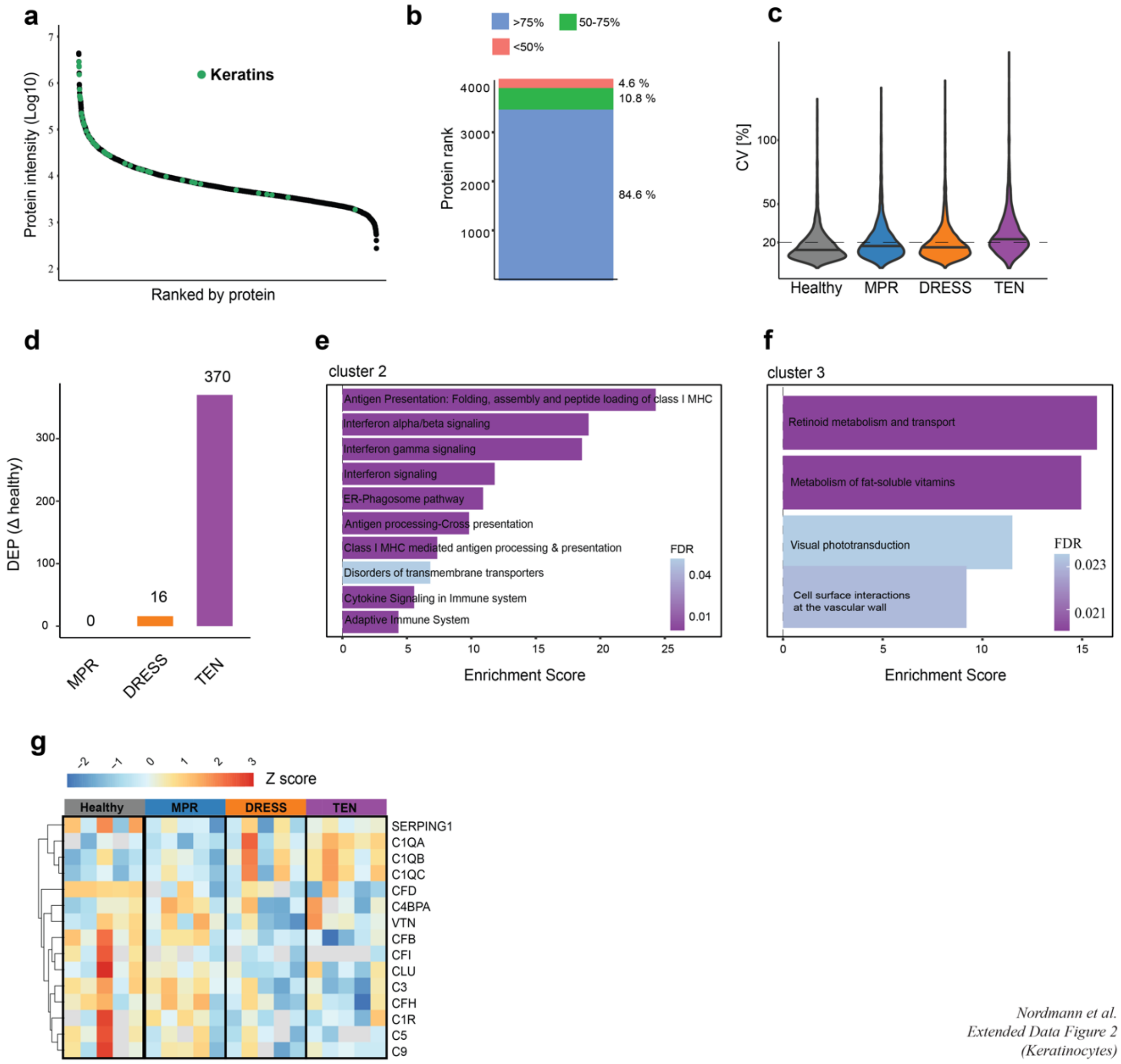


*Nordmann et al.*
*Extended Data Figure 2*
*(Keratinocytes)*

**Extended Data Fig. 2 (keratinocyte proteome) | a.** Abundance range rank plot of median protein intensity (y-axis). Keratins are highlighted in green. **b.** Data completeness for each protein across all samples binned into three groups (<50%, 50 – 75%, > 75%). **c.** Coefficient of variation (CV) of all proteins within the specified cohort. **d.** Number of differentially expressed proteins (DEP) compared to healthy. **e, f.** Overrepresentation analysis of ANOVA cluster 2 and 3 (Reactome), ordered according to enrichment score. Color indicates degree of significance (FDR). **g.** Semisupervised heatmap of complement factors in immune cells of the indicated conditions. Color indicates normalized intensity levels (z score). N = 5 individuals per cohort (TEN, DRESS, MPR and healthy).

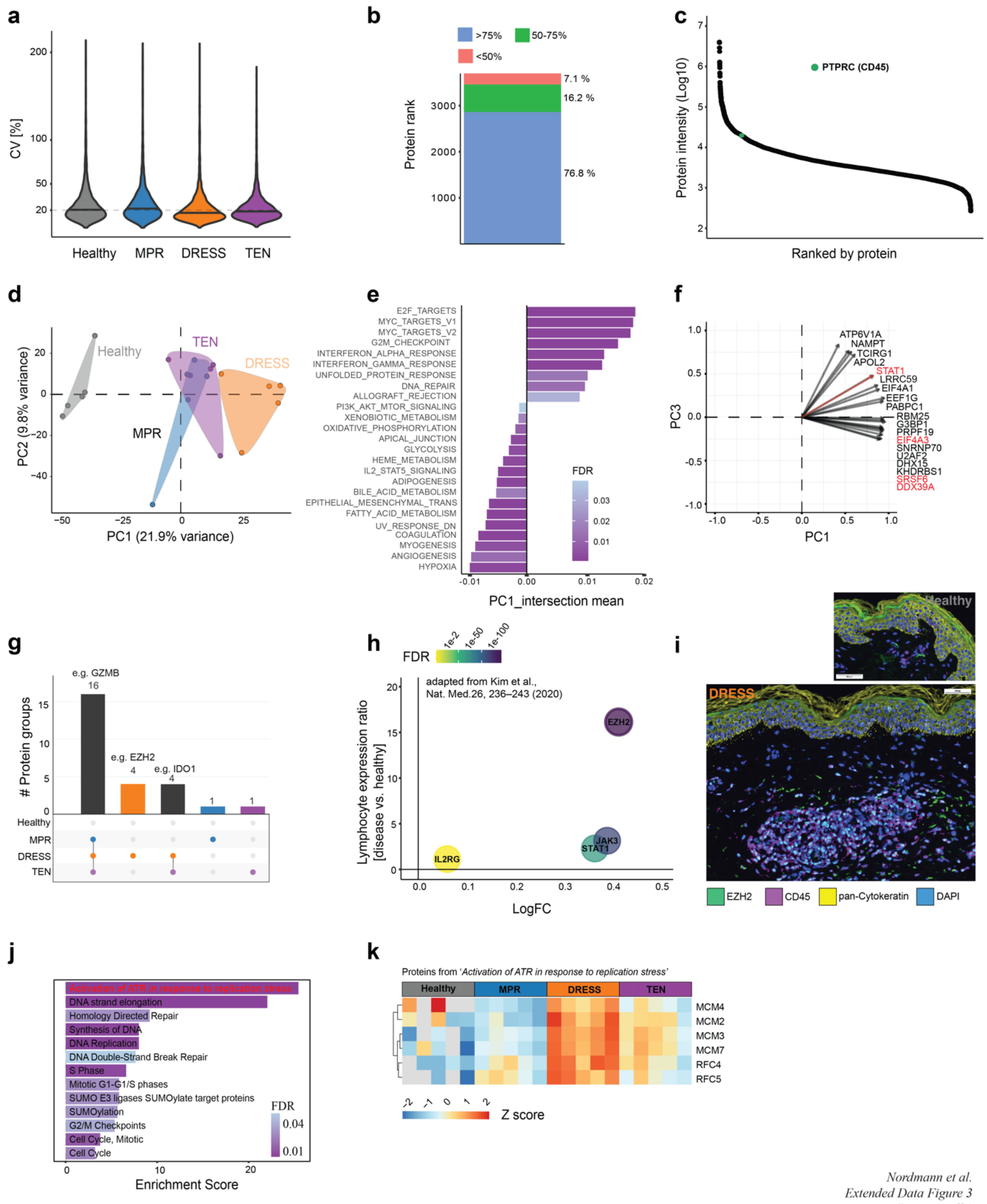


*Nordmann et al.*
*Extended Data Figure 3*
*(Immune cells)*

**Extended Data Fig. 3 (immune cell proteome) | a.** Coefficient of variation (CV) of all proteins within the specified cohort. **b.** Data completeness for each protein across all samples binned into three groups (<50%, 50 – 75%, > 75%). **c.** Abundance range rank plot of median protein intensity (y-axis). PTPRC as the segmentation marker of immune cells (CD45) is highlighted in green. **d.** Principal component analysis of PC1/2 **e.** PC1 gene set enrichment analysis (Hallmark) of immune cells, ordered according to enrichment score. Color indicates degree of significance (FDR). **f.** Correlation of top 20 proteins (loading factor) contributing to PC1/ 3. **g.** Upset plot of uniquely identified proteins specific to each of the indicated conditions. **h.** Comparative expression of

EZH2 and previously identified markers (STAT1, JAK3, IL2RG) in DRESS. Plot displays fold change (x-axis) and the ratio of cell expression in diseased versus healthy samples (y-axis) of differentially expressed genes in DRESS PBMC lymphocyte clusters. Source Data from Kim, D., Kobayashi, T., Voisin, B. et al. Targeted therapy guided by single-cell transcriptomic analysis in drug-induced hypersensitivity syndrome: a case report. Nat Med 26, 236–243 (2020). **i.** Representative immunofluorescent image of FFPE tissue sections stained for EZH2 (green), CD45 (purple), pan-Cytokeratin (yellow) and nuclear counterstain (blue) in DRESS and healthy. **j.** Overrepresentation analysis (Reactome) of ANOVA cluster 2, ordered according to enrichment score. Color indicates degree of significance (FDR). **k.** Semisupervised heatmap of enriched proteins of the most prominent term from Extended Data Fig. 3h, "activation of ATR in response to replication stress". Color indicates normalized intensities. N = 5 individuals per cohort (TEN, DRESS, MPR and healthy).

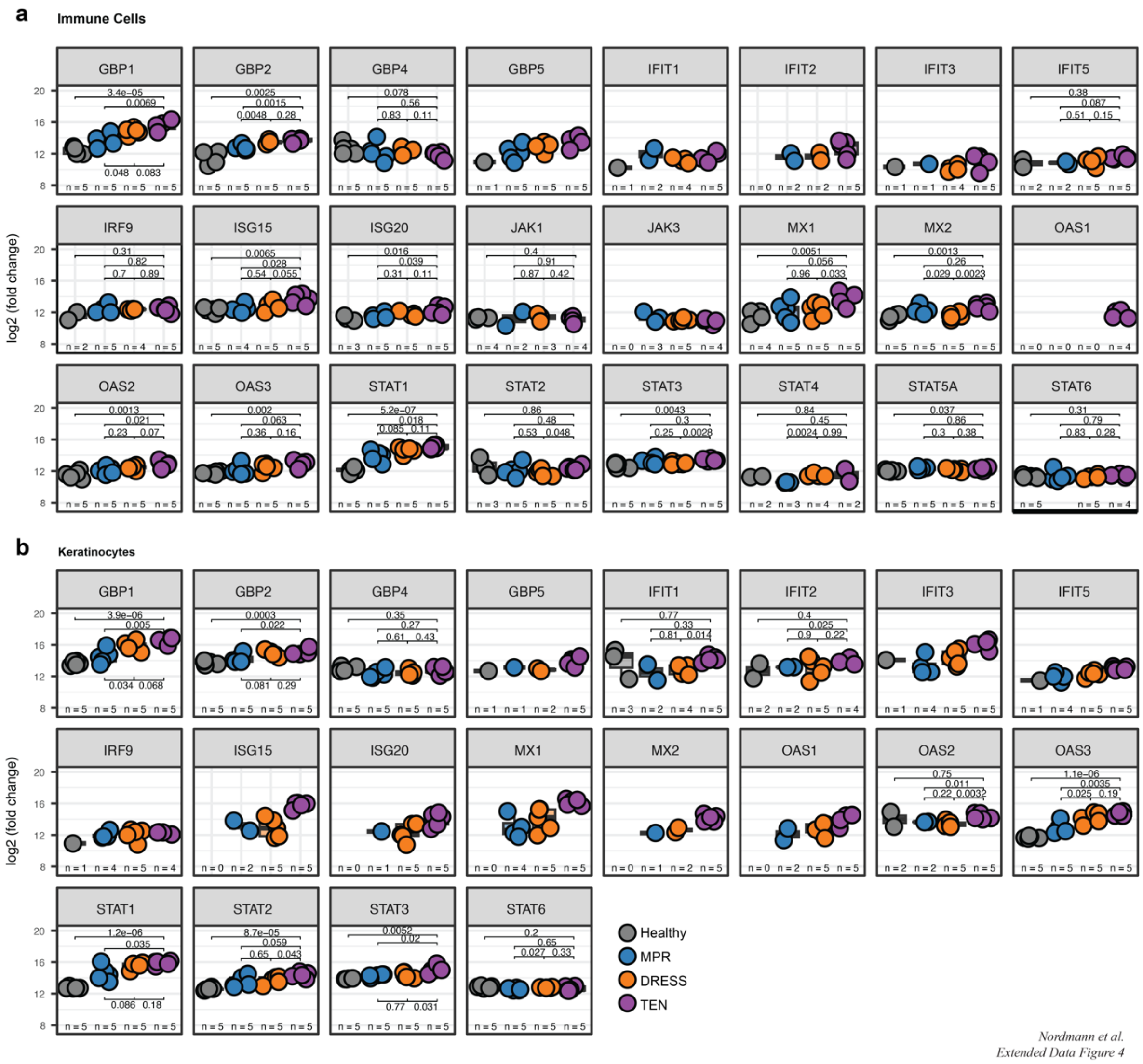


**Extended Data Fig. 4 | a,b** Comparative analysis of proteins involved in the JAK-STAT pathway (summarized in main Fig. 4b) across all cohorts in **a**, immune cells and **b**, keratinocytes (two-sided t-test). N = 5 individuals per cohort (TEN, DRESS, MPR and healthy). Box plots show the median (center line) with interquartile range of 25% to 75% and 95% confidence interval.

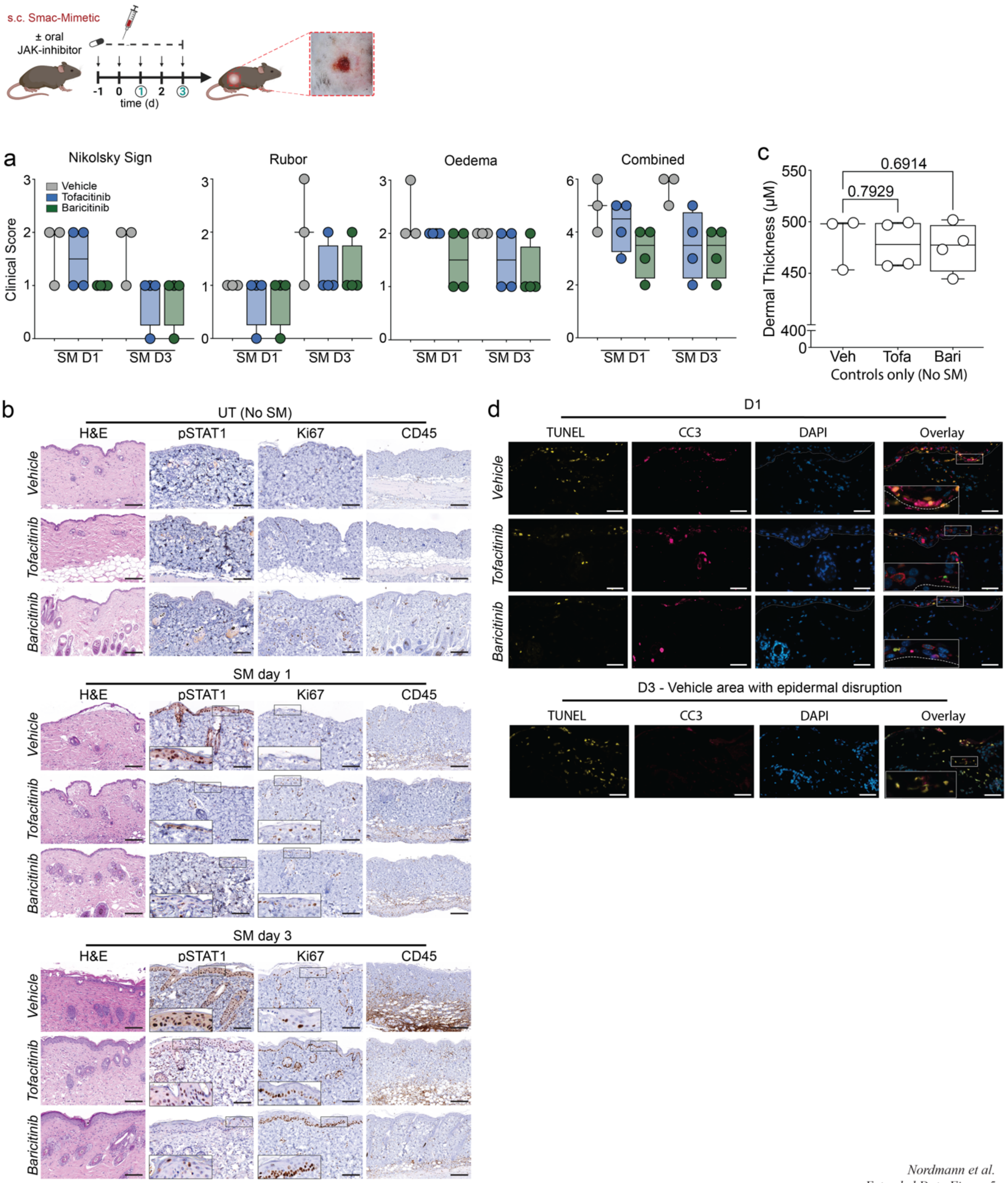


*Nordmann et al.*
*Extended Data Figure 5*

**Extended Data Fig. 5 | a.** Individual and combined clinical scores for the three assessment criteria (Epidermal disruption, Rubor, Oedema) evaluating clinical severity on days one (D1) and three (D3) after smac-mimetic (SM)-induced toxic epidermal necrolysis (TEN). **b** Representative H&E, pSTAT1, Ki67 and CD45 microscopy images, of the corresponding cohorts without SM (UT) and on D1 and D3 post TEN induction. Inserts magnify indicated area of interest. Scale bars = 100uM. **c.** Average dermal thickness of the control cohorts, without SM. Each data point represents 1 mouse, N=15 measurements/mouse. **d**. Immunofluorescence staining for cell death in the indicated cohorts following SM induced TEN. Magenta = cleaved caspase 3 (CC3, apoptosis), yellow = TUNEL (DNA fragmentation), cyan = DAPI (nucleus). Dotted lines indicate the dermo-epidermal junction. Box plots show the median (center line) with interquartile range of 25% to 75% and 95% confidence interval.

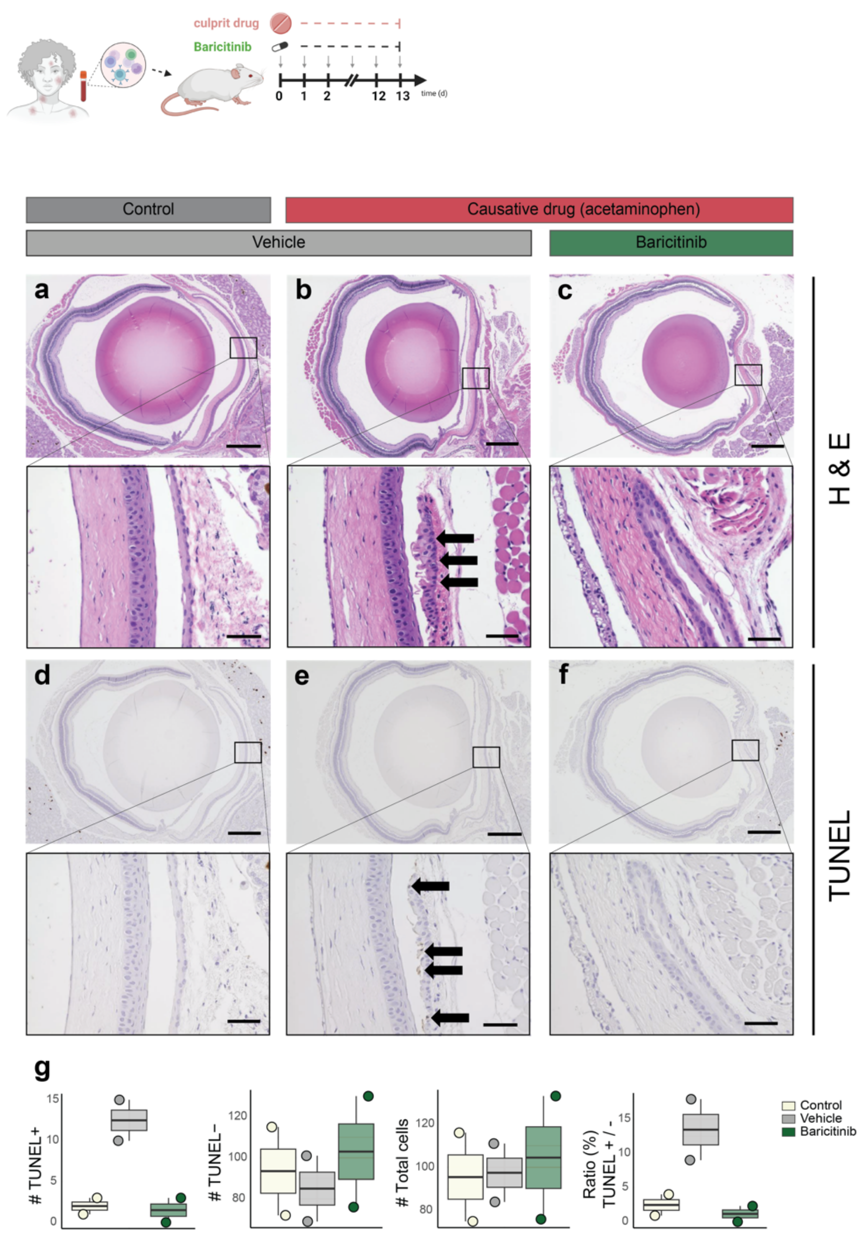


*Nordmann et al.*
*Extended Data Figure 6*

**Extended Data Fig. 6 | a - c,** Representative H&E and **d - f**, TUNEL immunohistochemistry images of the murine eye (scale bars = 500µm) across the cohorts. Inserts magnify the conjunctival subepithelial area of interest (scale bars = 50µm). Arrows indicate epithelial cell death (TUNEL$^{+}$). **g.** Quantification of TUNEL positive, TUNEL negative, their ratio and total count of subepithelial cells of the corresponding cohort. N = 2 mice per condition. Box plots show the median (center line) with interquartile range of 25% to 75% and 95% confidence interval.

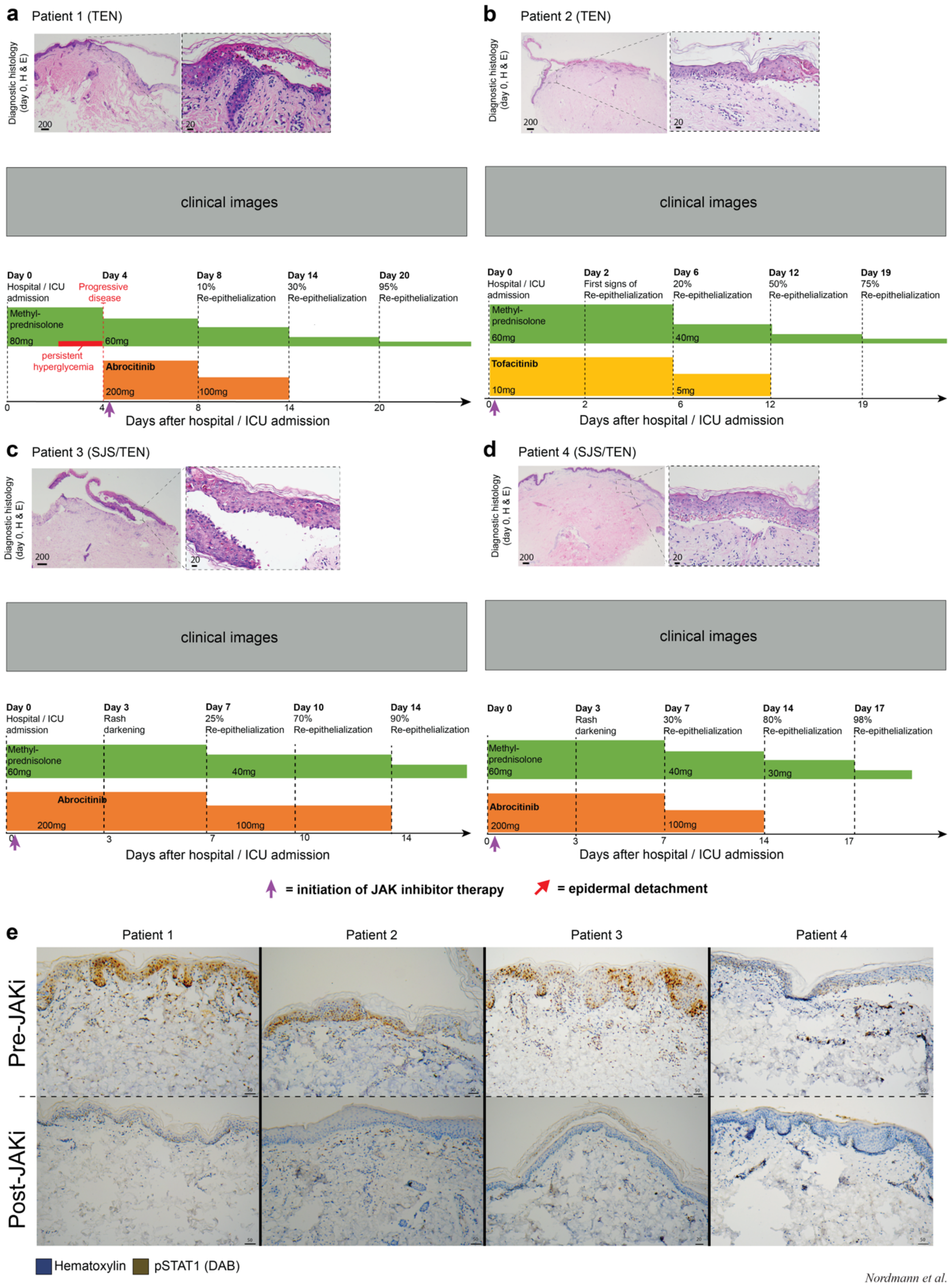


*Nordmann et al.*
*Extended Data Figure 7*

**Extended Data Fig. 7 | a.** as described in Fig. 6. **b.** patient with hepatocellular carcinoma under checkpoint inhibitor (Tislelizumab) therapy with clinically and histologically confirmed toxic epidermal necrolysis (TBSA 30 %, SCORTEN 3, predicted mortality 35.3 %) unresponsive to methylprednisolone (1mg/kg/day) monotherapy was given tofacitinib (200mg/day for seven days, 100mg/day for seven days). Rapid cessation of disease progression followed by 20 % re-epithelialization of detached skin areas six days later. **c.** patient with clinically and histologically confirmed Stevens-Johnson/TEN overlap syndrome (TBSA 10 %, SCORTEN 2, predicted mortality 12.1 %) associated with allopurinol intake. Upon admission she was immediately treated with methylprednisolone (1mg/kg/day) and abrocitinib (200mg/day for seven days, 100mg/day for seven days) resulting in cessation of progression with regression of erythema within 3 days and 25 % re-epithelialization of detached skin areas within seven days. **d.** patient with clinically and histologically confirmed Stevens-Johnson/TEN overlap syndrome (TBSA 12 %, SCORTEN 1, predicted mortality 3.2 %) associated with allopurinol intake. Upon admission, the patient was treated with methylprednisolone (1mg/kg/day) and abrocitinib (200mg/day for seven days, 100mg/day for seven days), resulting in cessation of progression with regression of erythema within 3 days and 30 % re-epithelialization of detached skin areas by day seven. All H&E stained FFPE lesional skin biopsies were taken on day 0 (= timepoint of hospital admission). **e.** pSTAT1 staining before and after JAK-inhibitor treatment, per patient. TBSA, total body surface area. Scale bars as indicated [μm].
